# Deconvoluting synovial fluid molecular endotypes in knee osteoarthritis: primary results from the STEpUP OA Consortium

**DOI:** 10.1101/2024.06.05.24308485

**Authors:** T.A. Perry, Y. Deng, P. Hulley, R.A. Maciewicz, J. Mitchelmore, S. Larsson, J. Gogain, S. Brachat, A. Struglics, C.T. Appleton, S. Kluzek, N.K. Arden, A.J. Price, D. Felson, L. Bondi, M. Kapoor, L.S. Lohmander, T.J. Welting, D.A. Walsh, A.M. Valdes, the STEpUP OA Consortium, L. Jostins-Dean, F.E. Watt, B.D.M. Tom, T.L. Vincent

## Abstract

**Background:** Osteoarthritis (OA) has a lifetime risk of over 40%, imposing a huge societal burden. Clinical variability suggests that it could be more than one disease. <u>S</u>ynovial fluid <u>T</u>o detect <u>E</u>ndoty<u>p</u>es by <u>U</u>nbiased <u>P</u>roteomics in OA (STEpUP OA) was established to test the hypothesis that there are detectable distinct molecular endotypes in knee OA.

**Methods:** OA knee synovial fluid (SF) samples (N=1361) were from pre-existing OA cohorts with cross-sectional clinical (radiographic and pain) data. Samples were divided into Discovery (N = 708) and Replication (N=653) datasets. Proteomic analysis was performed using SomaScan V4.1 assay (6596 proteins). Unsupervised clustering was performed using k-means, assessed using the f(k) metric, with and without adjustments for potential confounders. Regression analyses were used to assess protein associations with radiographic (Kellgren and Lawrence) and knee pain (WOMAC pain), with and without stratification by body mass index (BMI) or biological sex. Adjustments were made for cohort (random intercept) or intracellular protein, using an intracellular protein score (IPS). Analyses were carried out in R according to a pre-published plan.

**Results:** No distinct SF molecular endotypes were identified in OA but two indistinct clusters were defined in non-IPS regressed data which were stable across subgroup analyses. Clustering was lost after IPS regression adjustment. Strong, replicable protein associations were observed with radiographic disease severity, which were retained after adjustment for cohort or IPS. Pathway analysis identified a strong “epithelial to mesenchymal transition (EMT)” pathway, and weaker associations with “angiogenesis”, “complement” and “coagulation”. The latter were variably lost after adjustment for BMI or biological sex. Associations with patient reported pain were weaker.

**Conclusion:** These data support knee OA as a biologically continuous disease in which disease severity is associated with a strong, robust, tissue remodelling signature. Subtle differences were found in pathways after stratification by BMI or sex.

## BACKGROUND

Osteoarthritis (OA) of the knee is common, affecting up to a third of adults aged 60 years or older^[1]^. Characterised by failure of the synovial joint, OA is a major contributor to healthcare costs and is a leading cause of disability, manifesting as a spectrum of symptoms including chronic pain and limitations in function. Age and obesity are important risk factors, both of which have contributed to increasing disease burden across global populations^[2–4]^. There are currently no approved treatments for knee OA that effectively target structural disease and those that target symptomatic disease have modest efficacy and are associated with adverse events^[5, 6]^. There remains, therefore, a major unmet clinical need.

Limited understanding of disease pathogenesis coupled with a failure to translate findings from basic research to clinical settings has hampered clinical translation in OA^[7, 8]^. Another significant challenge is the broad clinical spectrum of disease that has led many to question whether OA is one disease, or whether it is driven by multiple different pathways that converge on a common joint pathology^[9, 10]^. Multiple clinical phenotypes have been suggested in the literature^[11–13]^, but these have not been validated as clinically useful stratification tools either when testing treatment responses or as predictors of disease progression^[14–16]^. Endotypes, defined by distinct molecular signatures, may have higher value, and could in part explain observable characteristics of a phenotype^[17]^.

Recent advances in understanding complex disease have been greatly enhanced by the application of multi-omic approaches to disease relevant tissues^[11, 18]^. The strengths of these approaches are the focus on human disease cohorts at scale (hundreds to tens of thousands of participating individuals), the unbiased and systematic nature of molecular identification, the ability to map molecules to a shared pathway, and the ability to replicate results across independent cohorts. Technological advances in genomics, transcriptomics and proteomics have enabled such studies to be carried out with low tissue volumes and at an affordable cost.

To date, the majority of studies that have attempted to identify molecular subgroups in OA have used systemic samples derived from blood (serum or plasma)^[19–21]^. The synovial fluid (SF), in contrast, offers a promising alternative discovery tissue, as it has proximity to the diseased tissues of the joint and is enriched with locally derived biomolecules. Thus, it is likely to represent more accurately the severity of disease in that given joint. We have also previously shown that proteins regulated in knee OA or after knee injury, compared with healthy controls, are readily detected in the SF but correlate poorly in paired blood^[22–25]^. Furthermore, we have previously confirmed the utility of high scale protein measurements in SF using the SomaScan® platform (SomaLogic, Inc, Boulder, Colorado), an aptamer-based assay^[26, 27]^. The SomaScan® platform V4.1 measures over 6596 distinct human proteins.

The <u>S</u>ynovial fluid <u>T</u>o detect <u>E</u>ndoty<u>p</u>es by <u>U</u>nbiased <u>P</u>roteomics in OA (STEpUP OA) Consortium was established to test the primary hypothesis that there are detectable distinct molecular endotypes in knee OA. We set out to perform an unsupervised analysis of a single SF sample from 1361 individuals with established OA where cross-sectional clinical data were also available. The standardised protocol, which describes the cohorts in detail, and includes how we adjusted for pre-defined technical and other confounding factors is available elsewhere^[27]^. Here we present the primary analysis of STEpUP OA, in which we determine whether protein molecular endotypes exist in the SF of participants with established knee OA, and further explore the relationship between proteomic signatures and structural and symptomatic disease.

## METHODS

### Study Design principles

STEpUP OA is an international Consortium, set up to search for molecular endotypes in knee OA utilising existing demographic factors including age, biological sex (verified through assessing the correlation between clinician-reported sex and four established sex biomarkers (PSA, FSH, LH and beta HCG)), body mass index (BMI) and clinical data (harmonised patient reported knee pain measures and radiographic scores) as well as matched knee SF samples (Supplementary Table 1). STEpUP OA utilised data and samples from 17 cohorts, including N = 1780 SF samples from 1676 individuals with established knee OA (by x-ray or knee joint symptoms), at risk of knee OA (following acute knee injury), or from control samples (disease-free or inflammatory arthritis participants). All participants gave written informed consent with local (institution specific) ethical approvals in place. Following the QC procedure, 1361 samples were identified from unique participants with established OA^[27]^. Individual cohorts were assigned, *a priori*, into Discovery (N = 708) and Replication (N = 653) datasets (Supplementary Table 1). Most samples were spun after joint aspiration but appropriate correction was applied when unspun samples were included in analyses. Full details of the cohorts and their associated metadata, how SF was collected and processed prior to SomaScan analysis, as well as how we corrected for predefined technical and other confounders can be found in Deng *et al*. 2023^[27]^. The primary Discovery statistical analysis was pre-specified and cross-sectional (Data Analysis Plan, see link below).

Sample numbers and SOMAmers®^[28]^ in the presented experiments varied according to data availability and analysis performed.

### Analysis platform

All SF samples were analysed on the Discovery Plex V4.1 (SomaLogic, Inc, Boulder, Colorado); a high-throughput, aptamer-based proteomics assay designed for the simultaneous assessment of 7596 synthetic DNA slow off-rate modified aptamers (SOMAmers®) (7289 unique human targets)^[29]^. All SF samples were randomized and analysed as a single batch at SomaLogic’s laboratory in Boulder, Co, USA.

### Statistical Analysis

#### Quality Control of Proteomic Data

All proteomic data received from SomaLogic underwent pre-processing and quality control procedures as previously reported^[27]^. Briefly, raw data was standardised using a modified version of SomaLogic’s normalization pipeline and batch-effect correction, followed by removal of samples and aptamers of insufficient quality to produce our initial downstream dataset for future analyses. All statistical analyses were pre-specified and outlined in our data analysis plans (see below).

#### Unsupervised clustering for endotype detection

Dimension reduction on batch-corrected, log-transformed proteomic data was performed using unscaled Principal Component Analysis (PCA), with the top principal components explaining 80% variation. Unsupervised clustering was performed on the reduced feature space using k-means clustering with 10 sets of random starting values. We tested for the presence of significant clusters using the f(K) statistic^[30]^; with the f(K) statistic visualised across cluster numbers. Data were determined to be significantly clustered if, for any number of clusters K, f(K)<0.85 (*a priori* specified). Elbow plots were constructed to test the robustness of our findings. If the data were significantly clustered, we picked the optimal cluster number by majority vote across different clustering metrics (as implemented in the R package *NbClust*^[31]^, version: 3.0.1) for downstream analyses.

#### Data visualisation and presentation

Clustering structure was visualised using Principal Component (PC) plots and Uniform Manifold Approximation and Projection for Dimension Reduction (UMAP)^[32]^ plots.

#### Protein–clinical feature association testing

Associations between protein expression and clinical outcomes were modelled by fitting regression models for each SOMAmer separately, with clinical features set as the dependent variable and log-expression for each protein set as the independent variable. Linear, logistic or proportional odds ordinal regression models were fitted for continuous, binary or ordered categorical variable outcomes respectively. Residual diagnostics confirmed adequacy of model assumptions. Before fitting the models, protein expression values were transformed using natural logarithms and were standardized on a per protein basis (within Discovery, Replication and Combined datasets) by subtracting mean log protein abundance and then dividing by its standard deviation, to make the slopes comparable between models. The resulting beta estimates (from linear regression models) or log odds ratios (from logistic and ordinal models) can be interpreted respectively as either mean outcome change or log odds ratio per standard deviation change in the log protein abundance. Replication was defined as proteins that were significant at Benjamini-Hochberg adjusted p-value ≤ 0.05 in both Discovery and Replication datasets and with effects in the same direction.

All primary regression models were adjusted for age and biological sex (with the exception of biological sex-stratified analyses that were adjusted for age only, and regression models exploring associations with BMI, which were adjusted for biological sex and radiographic disease status). All analyses were batch corrected for spin-status (using the R function *ComBat*^[33, 34]^) and run in duplicate using either proteomic data that had undergone further regression adjustment for intracellular protein score (IPS)^[27]^ (‘IPS regressed’ analyses) or without (‘non-IPS regressed’). Association testing between IPS, that had been transformed using natural logarithms, and demographic, clinical and technical features was performed using regression modelling, with all analyses either non-adjusted or adjusted for cohort (as a random intercept). Volcano plots were generated to display associated proteins from the regression analyses, with the most strongly positively and negatively associating proteins labelled by their given SomaLogic protein target name. The most significantly associated proteins, ordered by their adjusted p-value, were labelled. A small number of proteins (non-IPS & COMBAT corrected for spin-status filtered list: N = 383, IPS & COMBAT corrected for spin-status filtered list: N = 375), had more than one detection SOMAmer on the platform. Where this was the case, only one SOMAmer was labelled on the volcano plot (i.e. the most significant based on ranked adjusted p-value). We also conducted interaction testing for associations between protein abundance and clinical features of disease. A protein abundance-by-biological sex interaction term was included to test explicitly whether biological sex modified the association between protein abundance and WOMAC knee pain. Similarly, a protein abundance-by-obesity status (a dichotomous variable, BMI ≥ 30) interaction term was included to examine if associations with advanced radiographic status were modulated by protein abundance differences above and below this clinically relevant BMI threshold. Pre-specified clinical outcomes used in association testing are listed in (Supplementary Table 1).

#### Pathway enrichment analysis

We tested for enrichment of associated proteins within pathways using gene sets taken from The Molecular Signatures Database (MSigDB, https://www.gsea-msigdb.org/gsea/msigdb); specifically, Hallmark, Gene Ontology (GO), Reactome, and Kyoto Encyclopaedia of Genes and Genomes (KEGG). All proteins were mapped to the corresponding gene set based on ‘EntrezGeneSymbol’, ‘Target’ or ‘EntrezGeneID’ variables provided by SomaLogic. Protein set enrichment testing was performed using the *fgsea*^[35]^ package in R (version: 1.28.0) to identify pathways whose genes were enriched for association with a given outcome. All proteins featured in the respective regression models were ranked by a ‘rank metric’ calculated as; rank metric = -log(p-values) * sign(beta estimate or log odds ratio per standard deviation). The sign function returns +1 if the estimate is positive, −1 if it is negative, and 0 if it is zero thereby capturing the direction of effect (whether the feature is upregulated or downregulated). Enrichment scores were calculated as the maximum value of the running sum and normalized relative to pathway size, resulting in Normalized Enrichment Scores (NES). Direction and magnitude of pathway enrichment for a given outcome (i.e. differential regulation of the pathway) was determined using the NES score; with positive values representing positively associated pathways whilst negative values represented negatively associated pathways. The *ggplot2*^[36]^ R package (version: 3.5.0) was used to draw bubble plots and visualise results.

Protein-protein interaction (PPI) networks were constructed using the Search Tool for the Retrieval of Interacting Genes/Proteins database (STRING version 11.5, https://string-db.org/). The filter condition was set as follows: network type selected; “full-STRING network”; confidence ≥ 0.2-0.4.

#### Statistical Significance

Pearson correlation and relevant p-values are given for both correlation testing and regression modelling. All analyses were carried out in R (version 4.3.2), unless otherwise stated (R Core Team. (2016). R: A Language and Environment for Statistical Computing. Vienna, Austria. Retrieved from https://www.R-project.org/). Statistical significance was defined using Benjamini-Hochberg^[37]^ corrected p-values adjusted for multiple testing, at a false discovery rate (FDR) of 5% (padj ≤ 0.05).

**Data Analysis Plan:** https://www.kennedy.ox.ac.uk/oacentre/stepup-oa

## RESULTS

### Endotype Detection in OA SF

To search for molecular endotypes in OA using SF protein profiles, the f(K) cluster metric was employed. We had previously reported that a large contributor of variance in the initial processed data (principal component 1, accounting for 48% of variance), was due to intracellular proteins^[27]^. Appreciating that the intracellular protein signature could obscure subtle clustering patterns within the data, we performed cluster analyses with and without regression adjustment for intracellular protein^[27]^, using an intracellular protein score (IPS) that correlated highly with principal component 1 (r = 0.94)^[27]^. Cluster analysis revealed 2 clusters that were evident within Discovery, Replication and Combined datasets for the non-IPS regressed analysis (Figure 1A, left panel). In contrast, no clusters were detected in the IPS-regressed dataset (Figure 1A, right panel). Visualisation of the proteomic data structure in two-dimensional space showed that the two clusters were indistinct and could be defined by dichotomising the continuous IPS, a feature that was lost after IPS regression (Figure 1B).

**Figure 1:**
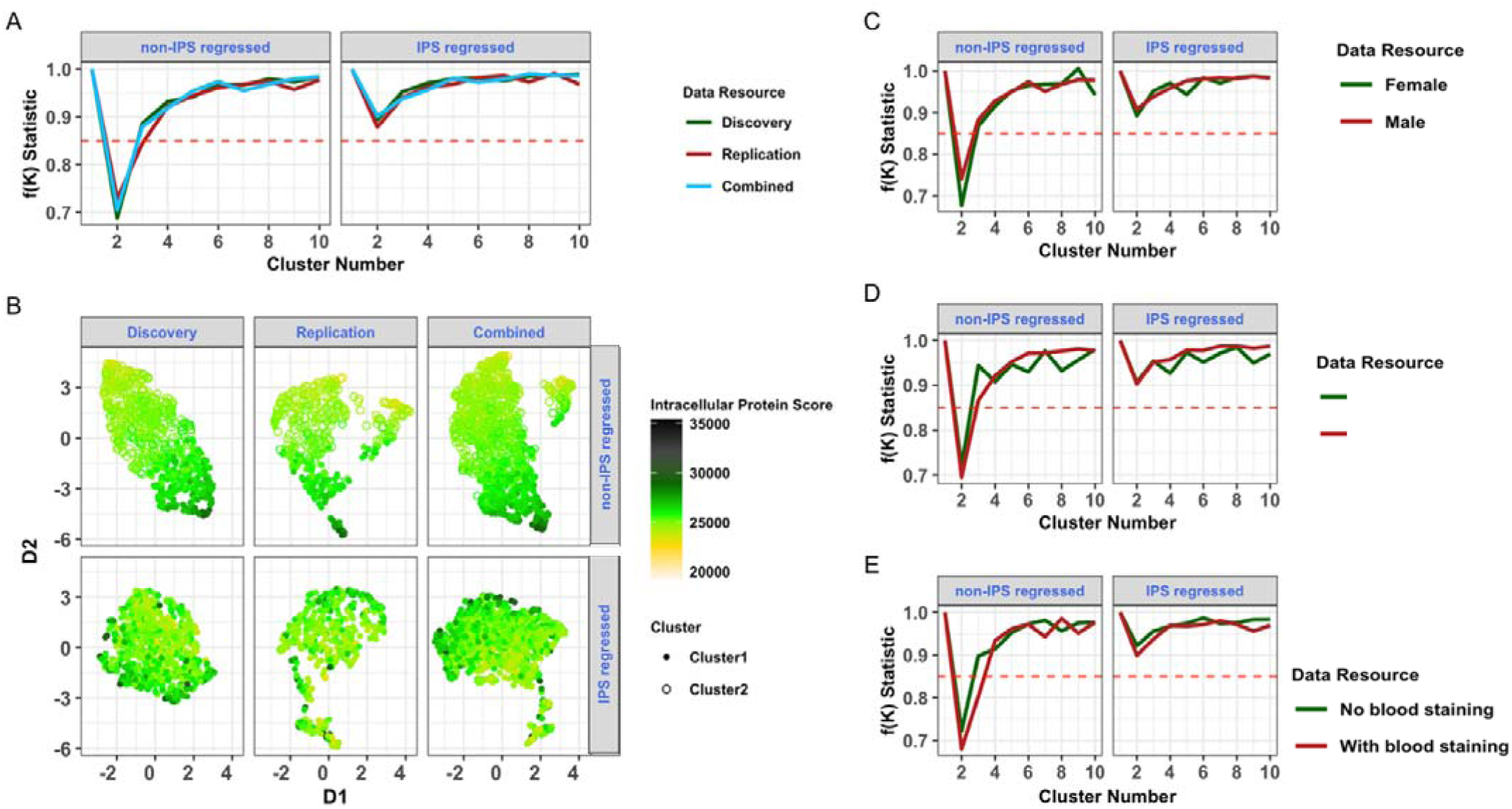
Endotype discovery by cluster analysis in Discovery, Replication and Combined datasets. **(A)** f(K) metric for non-IPS and IPS regressed analyses. Significant clustering was observed (f(k) < 0.85) within all three datasets for non-IPS regressed analyses only (left panel). **(B)** Visualisation of data structure and IPS on UMAP by dataset, stratified by non-IPS (top panel) and IPS regressed (bottom panel) analyses. f(K) metric plots for Combined dataset stratified by **(C)** biological sex, **(D)** radiographic disease severity (KL grades: 0-2 as ‘non advanced OA’ and ≥ 3 as ‘advanced OA’) or **(E)** blood staining (visual blood staining: 1 as ‘no blood staining’ and ≥ 2 as ‘with blood staining’) for non-IPS and IPS regressed analyses. Abbreviations: osteoarthritis (OA), intracellular protein score (IPS), Uniform Manifold Approximation and Projection (UMAP).

Association testing of IPS with pre-defined clinical and technical features (N = 1134, spun OA samples only) demonstrated that IPS was significantly, but modestly, greater in females, greater in advanced radiographic disease (KL grade ≥3), and was greater in SF samples with visual blood staining scores ≥2 (Table 1). We therefore repeated the cluster analysis, using IPS and non-IPS regressed datasets, but stratified by biological sex (Figure 1C), radiographic disease severity (Figure 1D) and presence of blood staining (Figure 1E). As with our non-stratified analyses, clusters (again indistinct) were only identified in non-IPS regressed data. Collectively these data suggest that there are two potential endotypes in the non-IPS corrected data, but they are on a continuum, defined by the IPS, and are not distinct. Furthermore, the cluster structure is independent of stage of disease, biological sex and visible blood staining.

**Table 1:**
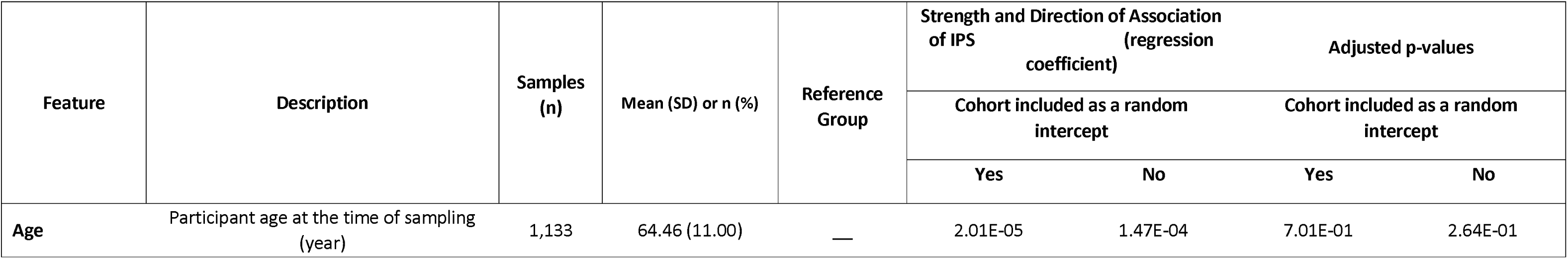
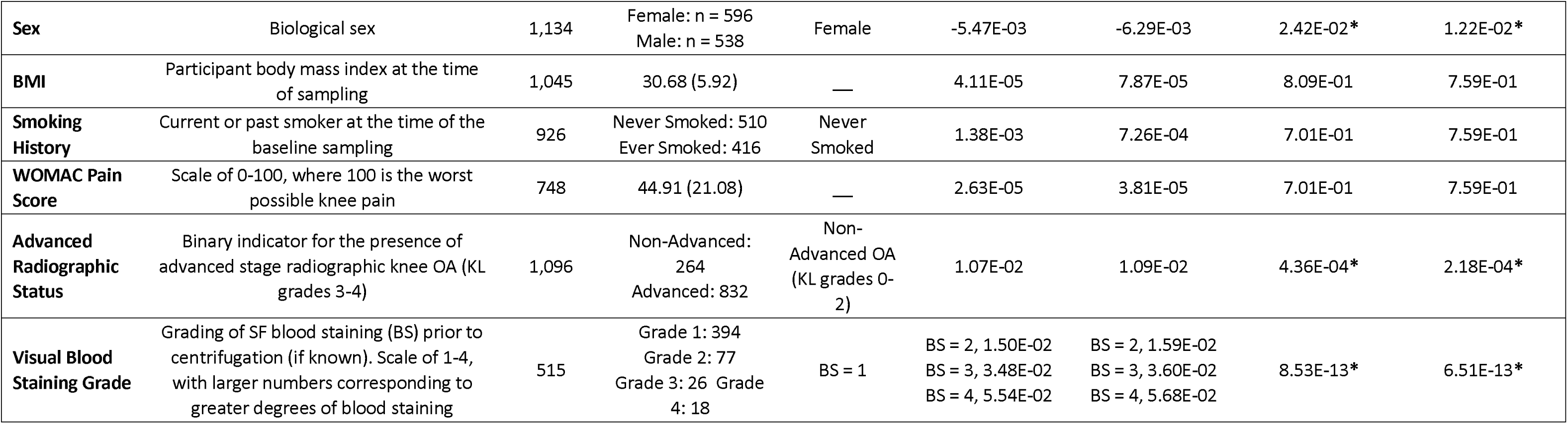
Baseline characteristics of participants, their SF samples and association of these factors with IPS. Association testing was carried out between IPS and core demographic, clinical and technical features in spun OA samples where relevant data were available. Linear regression models were constructed with log scaled IPS (i.e. IPS that were transformed using natural logarithms) as the outcome with each feature listed in the table used as a univariate exposure. Adjusted models where cohort was included as random intercept are also shown. Asterisks (bold) denote statistical significance at Benjamini-Hochberg cutoff (adjusted p-value ≤0.05). Abbreviations: osteoarthritis (OA), synovial fluid (SF), intracellular protein score (IPS), blood staining (BS), Kellgren Lawrence (KL), standard deviation (SD).

### Synovial Fluid protein associations with radiographic OA

We next examined which SF proteins were associated with radiographic disease severity. Over 1000 proteins were significantly associated with radiographic disease severity in each of the Discovery (N = 1021, 96.0% upregulated) and Replication datasets (N = 2524, 98.6% upregulated), with 688 (24.1%) proteins replicating across both datasets. Figure 2A shows the Combined dataset where 3815 proteins were associated with radiographic disease severity. Top associated proteins that replicated (across Discovery and Replication cohorts) and that remained significant in the Combined dataset after cohort adjustment, are labelled in orange. Protein abundance profiles for a selection of the labelled proteins were also significantly associated with ordinal KL grade, either significantly decreasing with worsening radiographic disease severity (LYVE1, IGFPB-6, FGFP1, sFRP-3) or increasing (TSG-6, sTREM-1, Activin A, VEGF121) (Figure 2B). Two additional proteins, associated with OA, MMP-13^[38]^ and COL2^[39]^, followed this latter pattern. Using the Hallmark gene set repository, nine differentially expressed pathways were significantly enriched across at least one of the three datasets (Figure 2C). Of these, “Epithelial Mesenchymal Transition (EMT)”, “Complement” and “Angiogenesis” were significantly associated with advanced radiographic OA across all datasets. Protein-protein interactions within each of the enriched pathways are shown in Figures 2D-F. “EMT” contained a number of molecules previously associated with matrix remodelling in OA^[40]^ including, but not limited to, TIMP1, TIMP3, MMP-2, TGFβ1, VEGFA and Fibronectin 1 (FN1). The correlation between protein associations within Discovery and Replication datasets was r = 0.49 (p<2.2 × 10^-16^) (Figure 2G).

**Figure 2:**
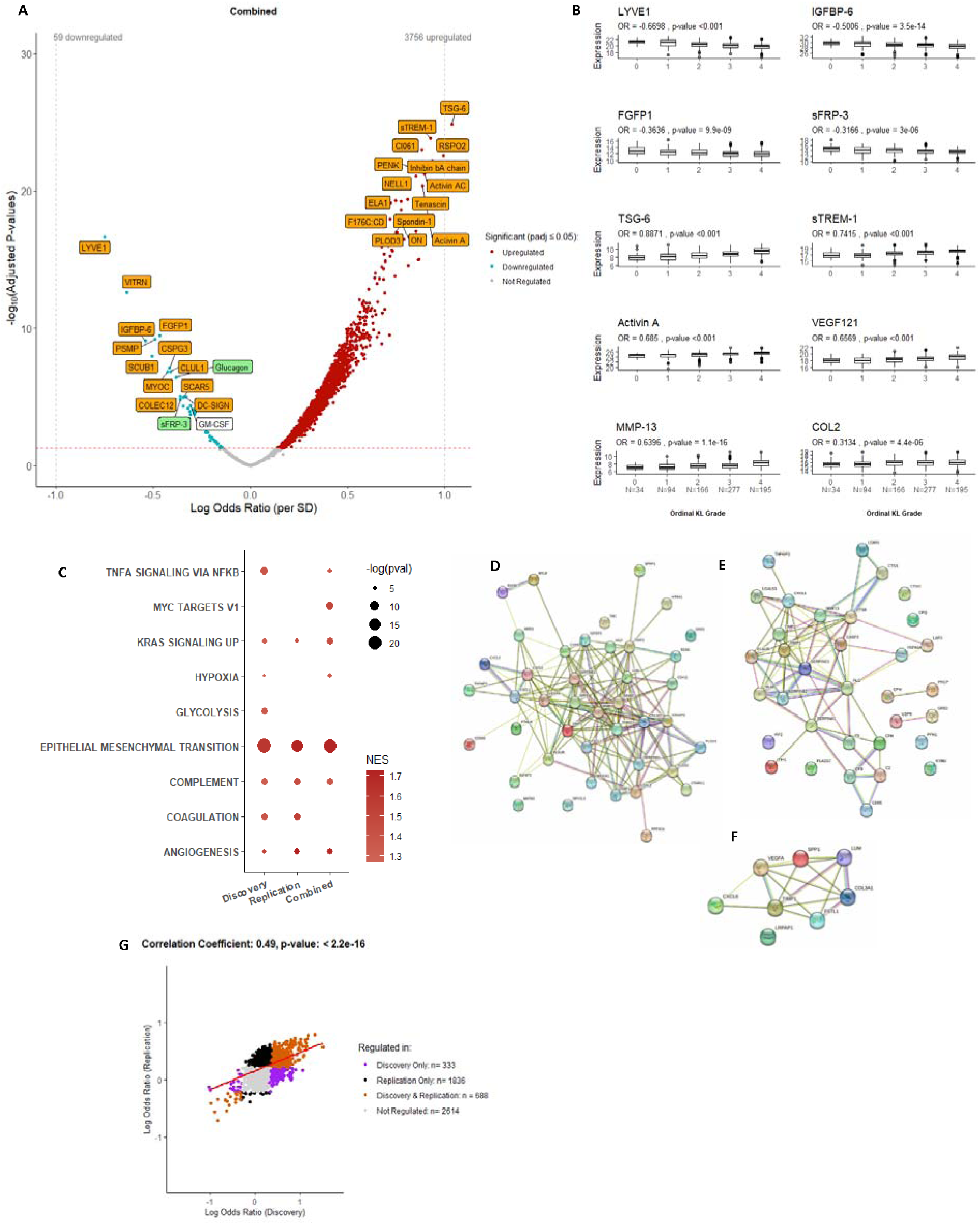
Association between protein abundance and advanced radiographic knee OA status in non-IPS regressed data. Protein abundance was measured in 1,322 samples (Discovery and Replication, spun (N=1,096) and unspun (N=226)), corrected for spin-status by ComBat and then adjusted for age and biological sex. **(A)** Volcano plot showing log odds ratios against adjusted p-values (Benjamini-Hochberg corrected) for proteins associated with advanced radiographic knee OA in the Combined dataset. Proteins in red are positively associated, those in blue negatively associated, with advanced radiographic status (KL grades: 3-4) at an adjusted p-value ≤ 0.05. Top associating proteins by adjusted p-value are labelled (top 30 positively and negatively associating proteins by adjusted p-value). Proteins that replicated (significant at padj ≤0.05 and with effects in the same direction in Discovery & Replication datasets), and that remained significant after the Combined dataset was adjusted for cohort (random intercept) are shown in orange. Proteins that either did not replicate but remained significant after adjustment for cohort, or did replicate but were not significant after cohort adjustment are shown in green. A single protein, GM-CSF neither replicated nor was significant after further adjustment for cohort (labelled white). **(B)** Select examples of protein expression values (transformed by natural logarithms) by ordinal KL grade (N=766) from the Combined dataset. Statistically significant associations with ordinal KL grade were tested by ordinal regression analysis (log odds ratio (OR) and unadjusted p-values are presented for each protein for models adjusted for age and biological sex). Two additional OA-related proteins (MMP-13 & COL2) are included. Number of samples in each group are shown. **(C)** Bubble plot of significantly enriched pathways (adjusted p-value <0.05) using the Hallmark Gene set for advanced RKOA status for Discovery, Replication and Combined non-IPS, non-cohort adjusted datasets. Protein-protein interaction networks, using STRING, for **(D)** Epithelial Mesenchymal Transition, **(E)** Complement and **(F)** Angiogenesis pathways. **(G)** Scatter plot of log odds ratio from logistic regression models of the associations between protein abundance and advanced radiographic disease status using either Discovery or Replication datasets is shown with significantly associated proteins in different datasets shown in different colours (see key). Pearson correlation coefficient and p-value (unadjusted) are presented for the correlation between log odds ratio generated in Discovery and Replication analyses. Abbreviations: intracellular protein score (IPS), Kellgren-Lawrence (KL). Lymphatic vessel endothelial hyaluronan receptor 1 (LYVE1), Insulin-like growth factor-binding protein 6 (IGFBP-6), Fibroblast Growth Factor Binding Protein 1 (FGFP1), secreted frizzled-related protein 3 (sFRP-3), tumour necrosis factor-inducible gene 6 (TSG-6), soluble triggering receptor expressed on myeloid cells-1 (sTREM-1), activin A and vascular endothelial growth factor A-(isoform 121)(VEGF-121), Matrix metalloproteinase-13 (MMP-13) and Collagen Type II (COL2), ON (osteonectin/SPARC). Full list of proteins available in Supplementary Data file 1.

We also performed similar analyses after correction for cohort (as a random intercept) or after IPS regression. Correlation of corresponding protein effects before and after cohort adjustment was high (r=0.88, p<2.2 × 10^-16^)(Supplementary Figure 1A), irrespective of differences in radiographic disease severity across cohorts (Supplementary Figure 1B). Pathway analysis showed a robust “EMT” signature across all datasets, although “complement” and “angiogenesis” pathways were no longer significantly enriched (Supplementary Figure 1C). For IPS regressed data, the volcano plot of proteins associated with radiographic disease severity is shown in Supplementary Figure 2A. Correlation of corresponding protein effects was also high (r=0.82, p<2.2 × 10^-16^)(Supplementary Figure 2B) and pathway associations for “EMT”, “complement” and “angiogenesis” remained robust, but also included “coagulation” (Supplementary Figure 2C). Data associated with these analyses can be found in Supplementary Data files 1 & 2.

### Synovial Fluid protein associations with advanced radiographic OA after stratification by BMI or biological sex

As “Metabolic OA”, driven largely by BMI, has been suggested as a potential OA phenotype^[41]^, we used STEpUP OA data to examine the proteins associated with radiographic disease severity after stratification by participant BMI (≥30 indicating obesity, N = 587 and <30, N = 649). We first looked at proteins in the SF that were associated with BMI, irrespective of disease status. Reassuringly, a number of proteins known to be associated with BMI, including the appetite suppressing hormone, leptin (LEP) insulin (INS), growth hormone receptor (GHR) and C-reactive protein (CRP) were identified (N = 248, 66.9% upregulated) (Supplementary Figure 3A; Supplementary Data file 3). Leptin’s SF levels correlated closely with BMI (r=0.58, p<2.2 × 10^-16^)(Supplementary Figure 3B) and associations of obesity-associated proteins appeared robust across datasets, and after cohort adjustment (Supplementary Figures 3C-E). When stratified by obesity status, over 1800 proteins were significantly associated with advanced radiographic OA in each of the obese and non-obese groups (Figure 3A, B), with a correlation between the corresponding protein effects in the obese and non-obese groups of r = 0.72 (p <2.2 × 10^-16^)(Figure 3C). No significant interaction terms with obesity status were identified by formal interaction testing (at padj <0.05). Interestingly, Hallmark pathway analysis showed a strong consistent “EMT” pathway signature in both groups, but only samples from obese participants retained significant associations with “coagulation” and “complement” (Figure 3D) (Supplementary Data file 4).

**Figure 3:**
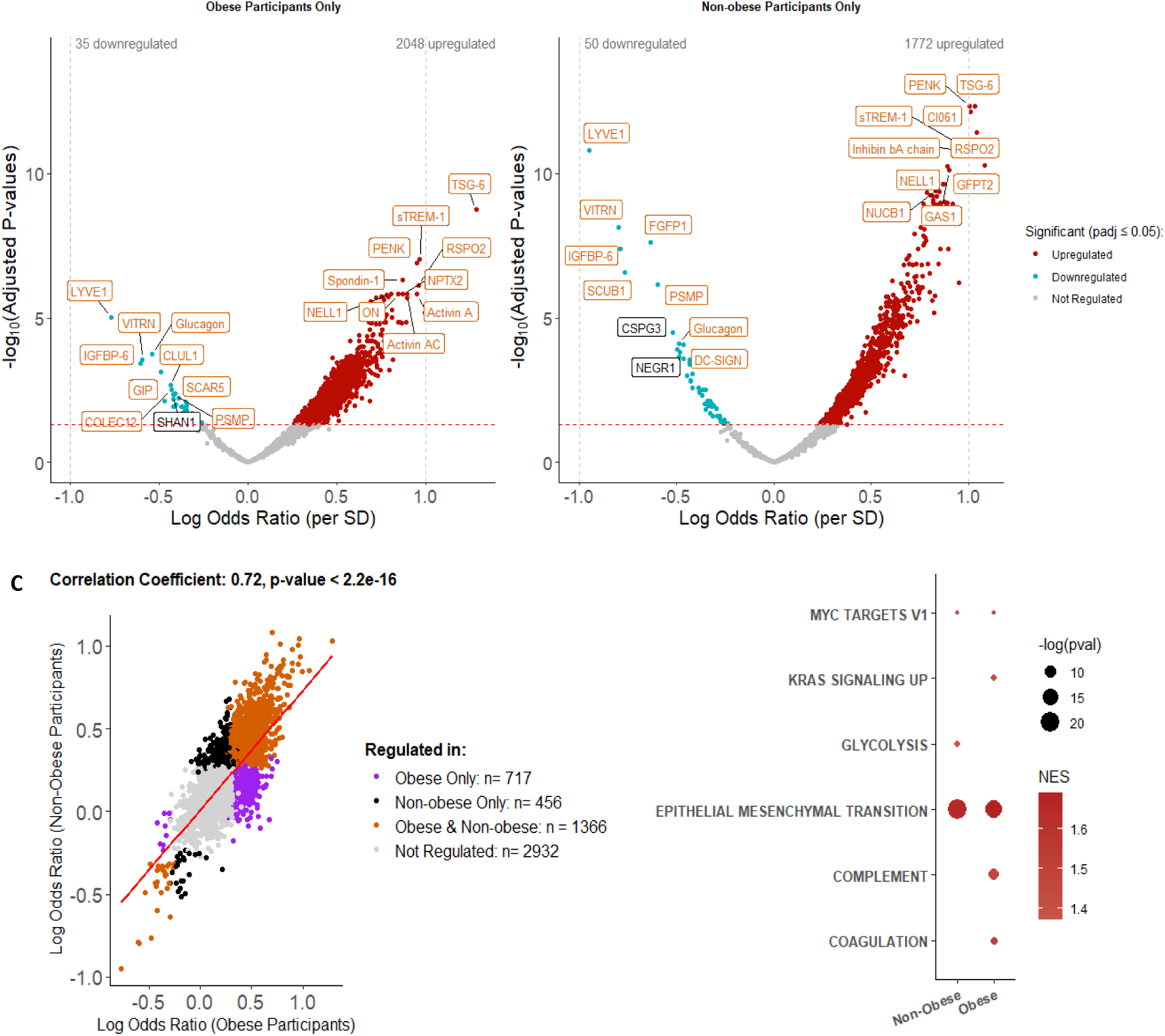
Association between protein abundance and advanced radiographic disease status stratified by obese and non-obese OA participants using non-IPS regressed Combined data. Protein abundance was measured in 1,236 patient samples where BMI was available (Combined dataset (N = 1,236, spun (N=1,045) and unspun (N = 191), corrected for spin-status and then adjusted for age and biological sex. The groups were then stratified by BMI into obese, BMI ≥30 (N = 587, 504 spun samples) and non-obese, BMI <30 (N = 649, 541 spun samples) participants. Volcano plots show log odds ratio against adjusted p-values (Benjamini-Hochberg corrected) for proteins associated with radiographic disease severity in **(A)** obese and **(B)** non-obese groups using Combined, non-IPS regressed and age adjusted data. Proteins in red are positively associated, those in blue negatively associated, with advanced radiographic disease at an adjusted p-value ≤ 0.05. Top 20 associated proteins by adjusted p-value are labelled. In orange are proteins that replicated (significant at padj ≤0.05 and with effects in the same direction) across obese and non-obese groups. **(C)** Pearson correlation of the log odds ratios comparing associations between protein expression and advanced radiographic disease status in obese and non-obese groups. **(D)** Bubble plot of significantly enriched pathways (adjusted p-value <0.05) using the Hallmark Gene set for proteins associated with advanced radiographic disease status by obesity status. Abbreviations: intracellular protein score (IPS). Full list of proteins is available in Supplementary data file 4.

To explore the influence of other participant factors on radiographic disease-protein associations, we also stratified samples by biological sex (Figure 4A, B). Protein associations with radiographic disease severity, after stratification by biological sex, also had a strong cross-strata correlation (r=0.69, p <2.2 × 10^-16^, Figure 4C), with 1437 significantly associated proteins common to the two groups. No significant interaction terms with biological sex were identified by formal interaction testing (at padj <0.05). Hallmark pathway analysis also showed a strong “EMT” pathway signature in both sexes, but only males showed significant associations with “angiogenesis” and “coagulation” (Figure 4D) (Supplementary Data file 5).

**Figure 4:**
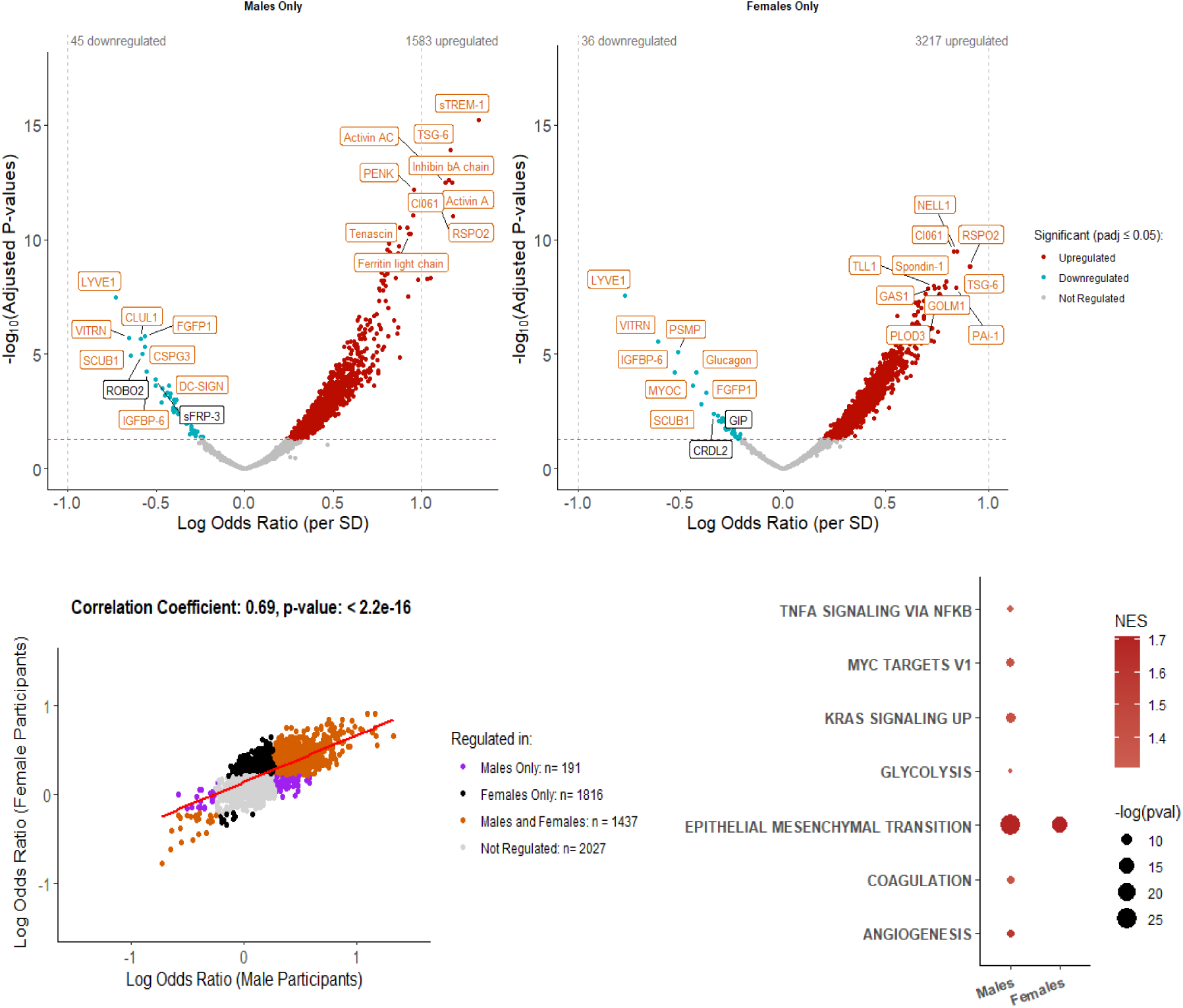
Association between protein abundance and radiographic OA severity after stratifying for biological sex using non-IPS regressed Combined data. Protein abundance was measured in 1,322 samples (Combined dataset, spun (N=1,096) and unspun (N = 226)), corrected for spin-status and then adjusted for age. Volcano plots showing odds ratios against adjusted p-values (Benjamini-Hochberg corrected) for proteins associated with advanced radiographic disease severity in the Combined dataset stratified by **(A)** males (N = 623) and **(B)** females (N = 699). Proteins in red are positively associated, those in blue negatively associated, with increased radiographic disease severity at an adjusted p-value ≤ 0.05. Top 20 associated proteins in each direction, by p-value, are labelled. Orange labelled proteins represent those that were significantly associated in both males and females, whereas white labelled proteins were only associated in the sex-specific set. **(C)** A scatter plot of log odds ratio from logistic regression models of the association between protein abundance and advanced radiographic disease status in males and females is shown with significantly associated proteins in different groups in different colours (see key). Pearson correlation coefficient and p-value (unadjusted) are presented for the correlation between log odds ratio generated in male and female sex-specific analyses. **(D)** Bubble plot of significantly enriched pathways (adjusted p-value <0.05) using the Hallmark Gene set for proteins associated with advanced radiographic disease status by biological sex. Abbreviations: intracellular protein score (IPS). Full list of proteins is available in Supplementary data file 5.

### Synovial Fluid protein associations with WOMAC pain in OA

Finally, we explored the association of SF proteins with patient reported pain. We identified 797 SF proteins that were significantly associated with WOMAC knee pain in the Combined non-IPS regressed dataset. However, none of these proteins replicated across Discovery and Replication datasets and the cross-dataset correlation was weak (r=0.36, p <2.2 × 10^-16^)(Figure 5A, B). Noelin-2 (NOE2) and ecto-ADP-ribosyltransferase 3 (NAR3) were the only significantly associated proteins in the Combined dataset after cohort adjustment (Supplementary Figure 4A and labelled green in Figure 5A). The relationships between NOE2 and NAR3 protein abundance with WOMAC pain subscores are shown in Figure 5C (Pearson correlation). The pathway analysis did not identify consistent associations across Discovery, Replication and Combined datasets (Figure 5D) and no significant pathways were identified within the Discovery dataset alone (at padj <0.05). Lack of replication may have been influenced by unevenly distributed knee pain subscores across Discovery and Replication cohorts (Supplementary Figure 4B). The number of proteins associated with pain was also reduced in the Combined dataset after adjustment for radiographic disease severity (Supplementary Figure 4C). NOE2 and NAR3 remained significantly associated with WOMAC pain after adjustment, and their levels were not independently associated with radiographic grade (by ordinal regression) (Supplementary Figure 4D). The correlation between pain-associated protein effects from non-IPS and IPS regressed analyses using the Combined datasets was r=0.97 (p <2.2 × 10^-16^) (Supplementary Figure 4E, Supplementary Data files 6 & 7). Further analyses on patient reported pain e.g. following stratification were not performed.

**Figure 5:**
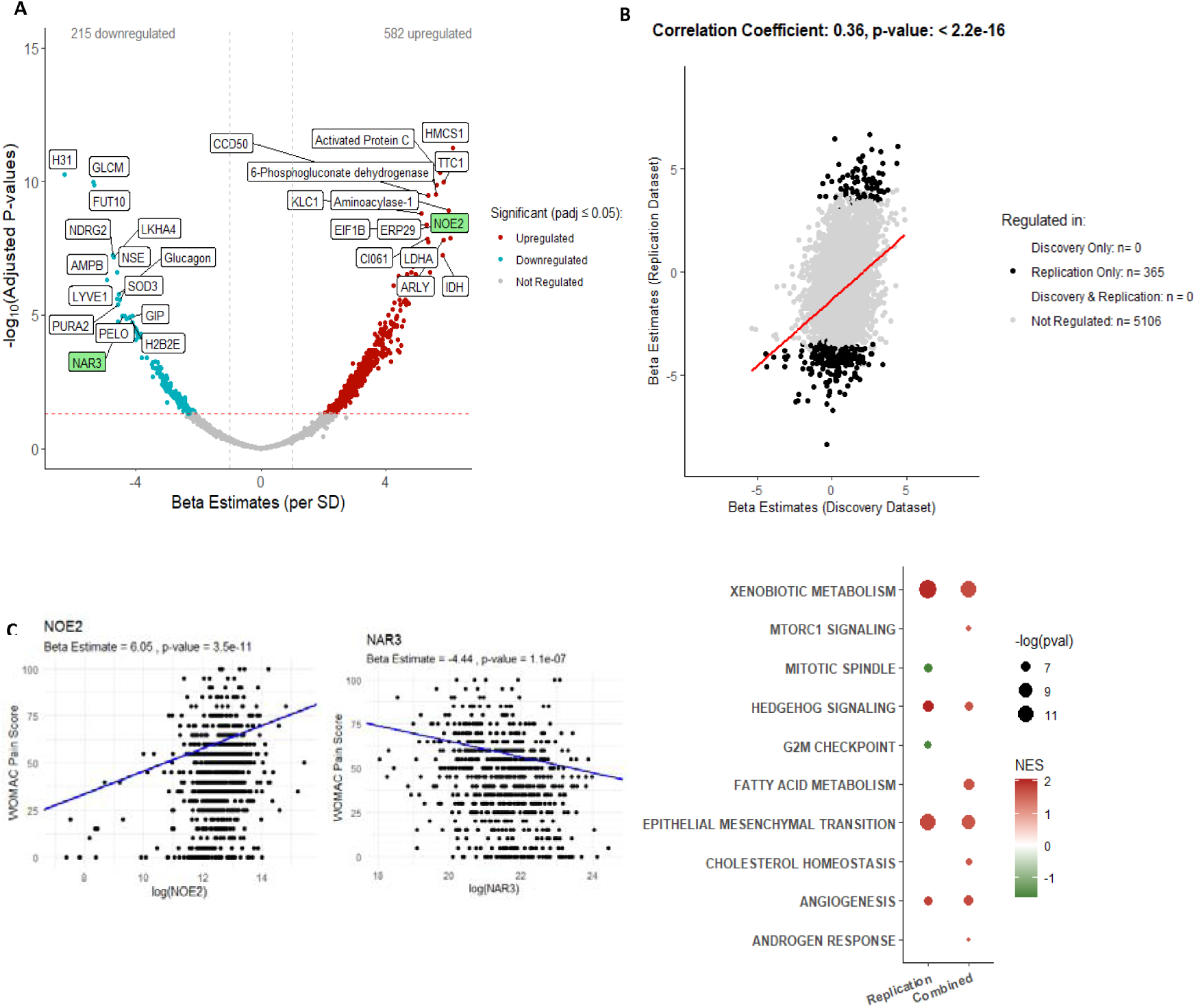
Association between protein abundance and WOMAC pain in non-IPS regressed data. Protein abundance was measured in 805 samples where WOMAC pain was available (Discovery and Replication, spun (N=748) and unspun (N = 57)), corrected for spin-status and then adjusted for age and biological sex. **(A)** Volcano plot showing beta estimates against adjusted p-values (Benjamini-Hochberg corrected) for proteins associated with WOMAC pain subscore in the Combined dataset. Proteins in red are positively associated, those in blue negatively associated, with increasing WOMAC pain subscore at an adjusted p-value ≤ 0.05. Top associated proteins, for each direction, ordered by adjusted p-value are labelled. In green are two proteins that were significant in Replication and Combined datasets, including after cohort adjustment (random intercept), see Supplementary Figure 4). **(B)** A scatter plot of beta estimates from linear regression models of the association between protein abundance and WOMAC knee pain in non-IPS analyses is shown for Discovery and Replication datasets with significantly associated proteins in different groups shown in different colours (see key). Pearson correlation coefficient and p-value (unadjusted) are presented for the correlation between beta estimates generated in non-IPS regressed analyses using Discovery and Replication datasets. **(C)** Scatter plots of WOMAC pain subscore against NOE2 or NAR3 protein abundance (transformed by natural logarithms) in OA participants using Combined, spin-status corrected, non-IPS regressed data. Beta estimates and p-values (unadjusted) are presented for linear models adjusted for age and biological sex. **(D)** Bubble plot of significantly enriched pathways (adjusted p-value <0.05) using the Hallmark Gene set for proteins associated with WOMAC knee pain by Replication and Combined datatsets not adjusted for IPS or cohort. No pathways were significantly enriched at padj <0.05 in the Discovery dataset. Abbreviations: osteoarthritis (OA), standard deviation (SD), intracellular protein score (IPS), Western Ontario and McMaster Universities Osteoarthritis Index (WOMAC, 0 = no pain, 100 = worst possible pain). Full list of proteins available in Supplementary data file 6.

## Discussion

In this manuscript we describe the primary results of STEpUP OA, the largest unbiased, replicated, cross-sectional synovial fluid proteomics analysis in knee OA ever performed. We uncover the balance of biological pathways in disease and how they change with structural and symptomatic disease severity. This dataset provides an unprecedented data resource from which to interrogate OA biology, address specific molecular questions and consider the influence of important patient-related factors, such as BMI and biological sex.

The data presented here do not reveal evidence for distinct molecular endotypes in knee OA SF, even when considering early radiographic disease separately. Rather, two continuous endotypes were identified by cluster analysis, which were defined by the IPS gradient. We still do not fully understand the importance or origin of intracellular protein in spun SF. Importantly, correcting for this signal using the IPS did not substantially change proteins or pathways associated with clinical features, suggesting that it is a minor influence on clinically relevant OA biology. It is therefore possible that the IPS-driven clustering is due to technical confounding during sample collection and processing. Taken together, the results support OA being a single heterogenous disease rather than multiple conditions each driven by a distinct pathway. This may appear at odds with studies suggesting discernible molecular clusters in tissues from participants with OA. Indeed, patient clusters have been described in the transcriptome of OA cartilage and synovium^[42–45]^, in SF using mass spectrometry^[46, 47]^, and in plasma^[17, 19, 21]^. However, these studies are smaller than STEpUP OA, and only a few included replication. Some of the studies examined prospective outcomes associated with clusters, rather than the cross-sectional analysis that we present here.

Synovial fluid is an ultrafiltrate of the plasma but also reflects joint-specific processes such as active secretion from cells^[48]^, including in extracellular vesicles, release from damaged or short-lived cells, and shedding from cell surfaces. Pathway analysis of knee OA SF proteins associated with radiographic disease severity indicates a robust activation of “EMT”, indicative of tissue remodelling, presumably part of the joint tissue injury response^[49]^. The “EMT” signature was consistent across all groups, irrespective of stratification and correction by cohort or IPS, or factors such as BMI and sex, suggesting that this is the common pathway in OA pathogenesis. Activation of complement, coagulation and angiogenesis was also evident, although was variable across subgroups. Whether these protein signatures identify groups of patients who display distinct treatment responses remains to be seen.

Replication across Discovery and Replication cohorts was robust for associations with structural disease but less so for pain. Patient reported outcome measures, such as knee pain, are known to be influenced by external factors beyond molecular drivers made by the joint e.g. psychological factors ^[50]^, making cross-sectional analyses of this sort challenging. Such extra-articular factors are complex and were not consistently collected within STEpUP OA cohorts. Protein associations with pain may also have been limited by the fact that WOMAC pain scores were only available on a subset within STEpUP OA (N = 805) and most of these were within a relatively narrow range of pain severity.

Despite this being the largest analysis of its kind in OA, we recognise a number of limitations: protein detection using the SomaScan platform, rather than mass spectrometry, is biased towards detection of full-length proteins, thus potentially missing fragments of proteins that could be biologically informative; our samples were generated from a diverse set of, largely, pre-existing cohorts and adjustment for cohort did reduce the number of significantly associated proteins; finally, by only focusing on proteins found in the synovial fluid, it is possible that key disease molecules or pathways were unintentionally excluded.

The cross-sectional analysis presented in this manuscript provides strong proof of concept that knee OA synovial fluid provides an informative window into disease-relevant biology. Future studies in STEpUP OA are now planned to ask whether SF signatures predict prospective clinical outcomes and whether they are driven in part by genetic variants associated with OA risk. Ultimately, we hope that SF analyses of this sort will assist in experimental medicine studies to test treatment responsiveness, helping to de-risk subsequent clinical trials of new interventions. The publication of this manuscript also marks the opportunity to welcome external parties to apply for access to STEpUP OA data for research purposes in accordance with our Consortium Agreement.

## Data Availability

The minimal datasets upon which this data relies and all R code, including the html vignette, are available at https://github.com/ndorms-tperry/STEpUP-OA-Primary-Manuscript. The full STEpUP OA dataset may be made available by application to the Data Access and Publication Group of STEpUP OA once the primary analysis manuscript is published, in accordance with what is stipulated in our Consortium Agreement. This may attract an access fee to cover administrative processing. Neither the minimal dataset nor the full STEpUP OA dataset include patient identifiable data.

## Author contributions

Conception and Design: TLV, FEW, LJD, PH, RAM, DP, SL, SB, LSL, AS, CTA, DF, BDMT, MK, TJW, DAW, AMV. Analysis and interpretation of data: TAP, YD, LJD, FEW, TLV, PH, RAM, JM, SB, BDMT, LB. Drafting Article: TLV, TAP, YD, LJD, FEW. Critical revision of article: all authors. Final Approval: all authors.

## Acknowledgements

We would like to express our gratitude and thanks to all cohort participants who contributed samples to STEpUP OA. We are grateful for the support from Floris Lafeber and Simon Mastbergen (Utrecht Medical Centre) for provision of samples. We thank the Oxford Knee Surgery Team. We thank Gretchen Brewer for her administrative support of the consortium.

## The STEpUP OA Consortium author block includes

University of Nottingham: Ana M. Valdes, David A. Walsh, Michael Doherty, Vasileios Georgopoulos; Lund University: Staffan Larsson, L. Stefan Lohmander, André Struglics; University of Cambridge: Brian D.M. Tom, Laura Bondi; University of Toronto: Mohit Kapoor, Rajiv Gandhi, Anthony Perruccio, Y. Raja Rampersaud, Kim Perry; University of Manchester: Tim Hardingham, David Felson; University of Oxford: Tonia L. Vincent, Thomas A. Perry, Luke Jostins-Dean, Yun Deng, Vicky Batchelor, Jennifer Mackay-Alderson, Gretchen Brewer, Rose M. Maciewicz, Brian Marsden, Nigel K. Arden, Philippa Hulley, Andrew Price, Stefan Kluzek, Megan Goff, Vinod Kumar, James Tey, Tamas Szommer; Imperial College London: Fiona E. Watt, Andrew Williams, Artemis Papadaki; University College Maastricht: Tim J. Welting, Pieter Emans, Tim Boymans, Liesbeth Jutten, Marjolein Caron, Guus van den Akker; University of Western Ontario: C. Thomas Appleton, Trevor B. Birmingham, J. Daniel Klapak; Biosplice: Sarah Kennedy, Jeymi Tambiah; Fidia: Devis Galesso, Nicola Giordan; SomaLogic: Joe Gogain, Darryl Perry, Anna Mitchel, Ela Zepko; Novartis: Sophie Brachat, Joanna Mitchelmore, Juerg Gasser, Lori Jennings; UCB: Waqar Ali.

## Supplementary Data

Supplementary files include:

Supplementary data files 1-7

## Patient and Public Involvement Statement

People with lived experience of osteoarthritis have been involved in the design of this project. A patient research panel was involved in discussing and inputting on the STEpUP OA project in February 2020 (invited to the Centre for Osteoarthritis Pathogenesis Versus Arthritis in Oxford, as part of its involvement activities). Aspects relevant to the development of the project were further discussed with the panel in July 2022. The working groups for the consortium included one focused on patient involvement and engagement. A lay summary is included in the appendix of our publicly available analysis plan. A short video about the project was produced and is available on our website: https://www.kennedy.ox.ac.uk/oacentre/stepup-oa/stepup-oa. In addition, the various constituent cohorts contributing to STEpUP OA also typically have lay or patient members on their steering committees.

## References

1. Neogi, T., The epidemiology and impact of pain in osteoarthritis. Osteoarthritis Cartilage, 2013. 21(9): p. 1145–53.

2. Safiri, S., et al., Global, regional and national burden of osteoarthritis 1990-2017: a systematic analysis of the Global Burden of Disease Study 2017. Ann Rheum Dis, 2020. 79(6): p. 819–828.

3. Morgan, O.J., et al., Osteoarthritis in England: Incidence Trends From National Health Service Hospital Episode Statistics. ACR Open Rheumatol, 2019. 1(8): p. 493–498.

4. Swain, S., et al., Trends in incidence and prevalence of osteoarthritis in the United Kingdom: findings from the Clinical Practice Research Datalink (CPRD). Osteoarthritis and Cartilage, 2020. 28(6): p. 792–801.

5. Karsdal, M.A., et al., Disease-modifying treatments for osteoarthritis (DMOADs) of the knee and hip: lessons learned from failures and opportunities for the future. Osteoarthritis Cartilage, 2016. 24(12): p. 2013–2021.

6. Oo, W.M., et al., The Development of Disease-Modifying Therapies for Osteoarthritis (DMOADs): The Evidence to Date. Drug Design Development and Therapy, 2021. 15: p. 2921–2945.

7. Makarczyk, M.J., et al., Current Models for Development of Disease-Modifying Osteoarthritis Drugs. Tissue Eng Part C Methods, 2021. 27(2): p. 124–138.

8. Cope, P.J., et al., Models of osteoarthritis: the good, the bad and the promising. Osteoarthritis Cartilage, 2019. 27(2): p. 230–239.

9. Deveza, L.A. and R.F. Loeser, Is osteoarthritis one disease or a collection of many? Rheumatology, 2018. 57: p. 34–42.

10. Hunter, D.J., Pharmacologic therapy for osteoarthritis-the era of disease modification. Nature Reviews Rheumatology, 2011. 7(1): p. 13–22.

11. Mobasheri, A., et al., The future of deep phenotyping in osteoarthritis: How can high throughput omics technologies advance our understanding of the cellular and molecular taxonomy of the disease? Osteoarthritis and Cartilage Open, 2021. 3(4): p. 100144.

12. Mobasheri, A., et al., Recent advances in understanding the phenotypes of osteoarthritis. F1000Res, 2019. 8.

13. Mobasheri, A., et al., Molecular taxonomy of osteoarthritis for patient stratification, disease management and drug development: biochemical markers associated with emerging clinical phenotypes and molecular endotypes. Curr Opin Rheumatol, 2019. 31(1): p. 80–89.

14. Deveza, L.A., A.E. Nelson, and R.F. Loeser, Phenotypes of osteoarthritis: current state and future implications. Clinical and experimental rheumatology, 2019. 37 Suppl 120 (5): p. 64–72.

15. Attur, M., et al., Prognostic biomarkers in osteoarthritis. Curr Opin Rheumatol, 2013. 25(1): p. 136–44.

16. Rocha, F.A.C. and S.A. Ali, Soluble biomarkers in osteoarthritis in 2022: year in review. Osteoarthritis Cartilage, 2023. 31(2): p. 167–176.

17. Luo, Y., et al., A low cartilage formation and repair endotype predicts radiographic progression of symptomatic knee osteoarthritis. J Orthop Traumatol, 2021. 22(1): p. 10.

18. Beier, F., The impact of omics research on our understanding of osteoarthritis and future treatments. Current Opinion in Rheumatology, 2023. 35(1): p. 55–60.

19. Angelini, F., et al., Osteoarthritis endotype discovery via clustering of biochemical marker data. Ann Rheum Dis, 2022. 81(5): p. 666–675.

20. Luo, Y.Y., et al., A low cartilage formation and repair endotype predicts radiographic progression of symptomatic knee osteoarthritis. Journal of Orthopaedics and Traumatology, 2021. 22(1).

21. Werdyani, S., et al., Endotypes of primary osteoarthritis identified by plasma metabolomics analysis. Rheumatology (Oxford), 2021. 60(6): p. 2735–2744.

22. Watt, F.E., et al., The molecular profile of synovial fluid changes upon joint distraction and is associated with clinical response in knee osteoarthritis. Osteoarthritis and Cartilage, 2020. 28(3): p. 324–333.

23. Watt, F.E., et al., Acute Molecular Changes in Synovial Fluid Following Human Knee Injury: Association With Early Clinical Outcomes. Arthritis Rheumatol, 2016. 68(9): p. 2129–40.

24. Struglics, A., et al., Changes in Cytokines and Aggrecan ARGS Neoepitope in Synovial Fluid and Serum and in C-Terminal Crosslinking Telopeptide of Type II Collagen and N-Terminal Crosslinking Telopeptide of Type I Collagen in Urine Over Five Years After Anterior Cruciate Ligament Rupture: An Exploratory Analysis in the Knee Anterior Cruciate Ligament, Nonsurgical Versus Surgical Treatment Trial. Arthritis & Rheumatology, 2015. 67(7): p. 1816–1825.

25. Garriga, C., et al., Clinical and molecular associations with outcomes at 2 years after acute knee injury: a longitudinal study in the Knee Injury Cohort at the Kennedy (KICK). Lancet Rheumatology, 2021. 3(9): p. E648–E658.

26. Broomfield, J.A.J., Using synovial fluid biomarkers to define a phenotype of osteoarthritis in the hip [PhD thesis]. 2020, University of Oxford.

27. Deng, Y., et al., *Methodological development of molecular endotype discovery from synovial fluid of individuals with knee osteoarthritis:* the STEpUP OA Consortium. medRxiv, 2023: p. 2023.08.14.23294059.

28. Rohloff, J., et al., Nucleic Acid Ligands With Protein-like Side Chains: Modified Aptamers and Their Use as Diagnostic and Therapeutic Agents. Molecular therapy. Nucleic acids, 2014. 3: p. e201.

29. Candia, J., et al., Assessment of variability in the plasma 7k SomaScan proteomics assay. Sci Rep, 2022. 12(1): p. 17147.

30. Pham, D.T., S.S. Dimov, and C.D. Nguyen, Selection of K in K-means clustering. Proceedings of the Institution of Mechanical Engineers Part C-Journal of Mechanical Engineering Science, 2005. 219(1): p. 103–119.

31. Charrad, M., et al., Nbclust: An R Package for Determining the Relevant Number of Clusters in a Data Set. Journal of Statistical Software, 2014. 61(6): p. 1–36.

32. McInnes, L. and J. Healy, *UMAP:* Uniform Manifold Approximation and Projection for Dimension Reduction. ArXiv, 2018. abs/1802.03426.

33. ComBat: Adjust for batch effects using an empirical Bayes framework. 2022 [cited10-02-2024]; Available from: https://rdrr.io/bioc/sva/man/ComBat.html.

34. Johnson, W.E., C. Li, and A. Rabinovic, Adjusting batch effects in microarray expression data using empirical Bayes methods. Biostatistics, 2007. 8(1): p. 118–127.

35. Korotkevich, G., et al., Fast gene set enrichment analysis. bioRxiv, 2021: p. 060012.

36. Valero-Mora, P.M., *ggplot2: Elegant Graphics for Data Analysis.* Journal of Statistical Software, Book Reviews, 2010. 35(1): p. 1–3.

37. Benjamini, Y. and Y. Hochberg, Controlling the False Discovery Rate: A Practical and Powerful Approach to Multiple Testing. Journal of the Royal Statistical Society. Series B (Methodological), 1995. 57(1): p. 289–300.

38. Wang, M., et al., MMP13 is a critical target gene during the progression of osteoarthritis. Arthritis research & therapy, 2013. 15(1): p. R5.

39. Garnero, P., et al., Cross sectional evaluation of biochemical markers of bone, cartilage, and synovial tissue metabolism in patients with knee osteoarthritis: relations with disease activity and joint damage. Annals of the rheumatic diseases, 2001. 60(6): p. 619–26.

40. Aigner, T., et al., Large-scale gene expression profiling reveals major pathogenetic pathways of cartilage degeneration in osteoarthritis. Arthritis and rheumatism, 2006. 54(11): p. 3533–44.

41. Zhang, W., et al., Classification of osteoarthritis phenotypes by metabolomics analysis. BMJ Open, 2014. 4(11): p. e006286.

42. Soul, J., et al., Stratification of knee osteoarthritis: two major patient subgroups identified by genome-wide expression analysis of articular cartilage. Ann Rheum Dis, 2018. 77(3): p. 423.

43. Steinberg, J., et al., Linking chondrocyte and synovial transcriptional profile to clinical phenotype in osteoarthritis. Ann Rheum Dis, 2021. 80(8): p. 1070–1074.

44. Wijesinghe, S.N., et al., Obesity defined molecular endotypes in the synovium of patients with osteoarthritis provides a rationale for therapeutic targeting of fibroblast subsets. Clin Transl Med, 2023. 13(4): p. e1232.

45. Fernandez-Tajes, J., et al., Genome-wide DNA methylation analysis of articular chondrocytes reveals a cluster of osteoarthritic patients. Ann Rheum Dis, 2014. 73(4): p. 668–77.

46. Ali, N., et al., Proteomics Profiling of Human Synovial Fluid Suggests Increased Protein Interplay in Early-Osteoarthritis (OA) That Is Lost in Late-Stage OA. Mol Cell Proteomics, 2022. 21(3): p. 100200.

47. Carlson, A.K., et al., Characterization of synovial fluid metabolomic phenotypes of cartilage morphological changes associated with osteoarthritis. Osteoarthritis Cartilage, 2019. 27(8): p. 1174–1184.

48. Timur, U.T., et al., Identification of tissue-dependent proteins in knee OA synovial fluid. Osteoarthritis Cartilage, 2021. 29(1): p. 124–133.

49. Muthu, S., et al., Failure of cartilage regeneration: emerging hypotheses and related therapeutic strategies. Nat Rev Rheumatol, 2023. 19(7): p. 403–416.

50. Bartley, E.J., S. Palit, and R. Staud, Predictors of Osteoarthritis Pain: the Importance of Resilience. Current rheumatology reports, 2017. 19(9): p. 57.

