## Supplementary Figures & Tables for "Deconvoluting synovial fluid molecular endotypes in knee osteoarthritis: primary results from the STEpUP OA Consortium"

|  | | | **STEpUP OA DATASETS** | | |
| --- | --- | --- | --- | --- | --- |
|  |  |  | **DISCOVERY** | **REPLICATION** | **COMBINED** |
|  |  |  | N = 1,045 total samples ^1^ | N = 701 total samples^1^ | N = 1,746 total samples^1^ |
|  |  |  | N = 719 baseline OA samples (***before*** *filtering*)^2^ | N = 664 baseline OA samples (***before*** *filtering*)^2^ | N = 1,383 baseline OA samples ***before*** *filtering*)^2^ |
|  | | | N = 708 baseline OA samples (***after*** *filtering*)^3^ | N = 653 baseline OA samples (***after*** *filtering*)^3^ | N = 1,361 baseline OA samples (***after*** *filtering*)^3^ |
| **Feature** | **Feature Description** | **Number of samples with available data (Discovery, Replication and Combined datasets)** | **Median (IQR) or n (%)** | | |
| **Age** | Participant age at the time of sampling (year) | 707, 653, 1360 | 65 (15) | 66 (14) | 66 (15) |
| **Sex** | Biological sex | 708, 653, 1361 | f = 366 (52%), m = 342 (48%) | f = 344 (53%), m = 309 (47%) | f = 710 (52%), m = 651 (48%) |
| **BMI** | Participant body mass index at the time of sampling | 694, 542, 1236 | 29.61 (7.36) | 29.72 (7.68) | 29.67 (7.55) |
| **Smoking History** | Current or past smoker at the time of the baseline sampling | 623, 477, 1100 | Never smoked: n = 332 (53%)  Ever smoked: n = 291 (47%) | Never smoked: n = 267 (56%)  Ever smoked: n = 210 (44%) | Never smoked: n = 599 (55%)  Ever smoked: n = 501 (45%) |
| **WOMAC Pain Score** | Scale of 0-100, where 100 is the worst possible knee pain | 420, 385, 805 | 50 (25) | 40 (40) | 45 (30) |
| **Advanced Radiographic Status** | Binary indicator for the presence of advanced stage radiographic knee OA (KL grades 3-4) | 705, 617, 1322 | Non-advanced: n = 131 (19%)  Advanced: n = 574 (81%) | Non-advanced: n = 175 (28%)  Advanced: n = 445 (72%) | Non-advanced: n = 306 (23%)  Advanced: n = 1016 (77%) |
| **Ordinal KL Grade** | Kellgren Lawrence grade (0-4)  (worst affected compartment) | 187, 579, 766 | 0: 19 (10%)  1: 35 (19%)  2: 65 (35%)  3: 43 (23%)  4: 25 (13%) | 0: 15 (3%)  1: 59 (10%)  2: 101 (18%)  3: 234 (40%)  4: 170 (29%) | 0: 34 (4%)  1: 94 (12%)  2: 166 (22%)  3: 277 (36%)  4: 195 (26%) |
| **Spin Status** | Binary indicator for whether SF sample had been centrifuged prior to supernatant storage | 708, 653, 1361 | Unspun: n = 1 (0.1%)  Spun: n = 707 (99.9%) | Unspun: n = 226 (35%)  Spun: n = 427 (65%) | Unspun: n = 227 (17%)  Spun: n = 1134 (83%) |
| ^1^ Total number of individual synovial fluid samples that passed the quality control (QC) procedure (Deng *et al*. 2023) before additional QC sample filtering.  ^2^ Total number of individual baseline (first available visit) OA synovial fluid samples before additional QC sample filtering.  ^3^ Total number of individual baseline OA synovial fluid samples after additional QC sample filtering (N = 11 samples were removed from both Discovery & Replication) – samples of poor quality were removed. | | | | | |

**Supplementary Table 1: *Characteristics of Discovery, Replication and Combined datasets and participant samples.*** Demographic and clinical data are summarized. Abbreviations: female (f), male (m), Kellgren Lawrence (KL), Western Ontario and McMaster Universities Osteoarthritis Index (WOMAC), interquartile range (IQR), radiographic knee osteoarthritis (OA) (RKOA), quality control (QC), metres (m), kilograms (kg), body mass index (BMI), synovial fluid (SF).

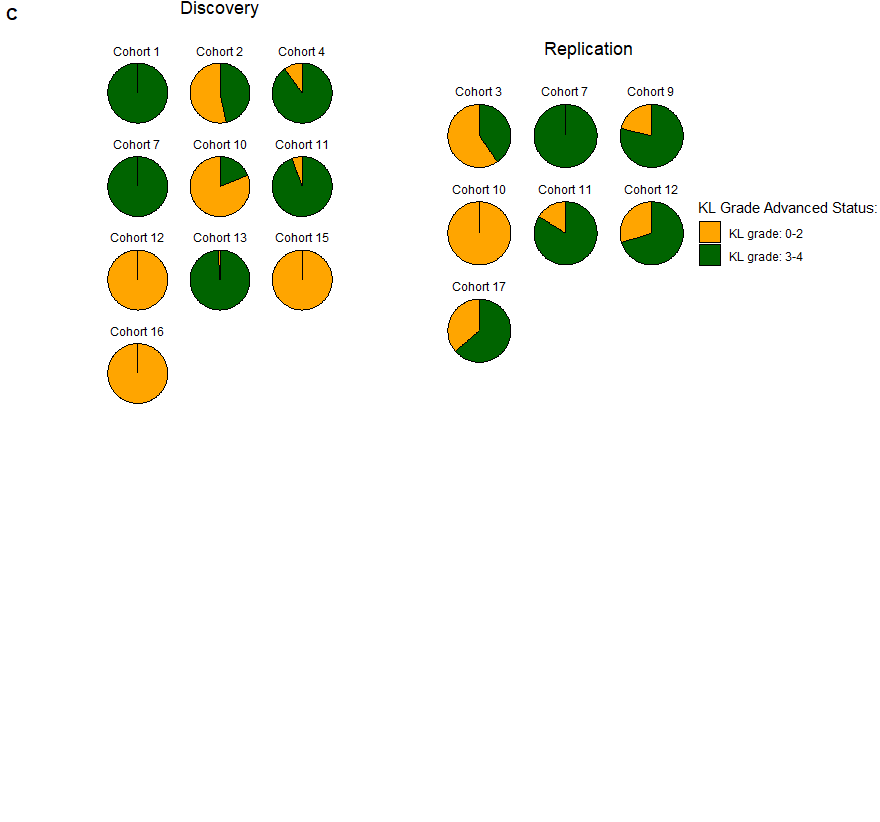

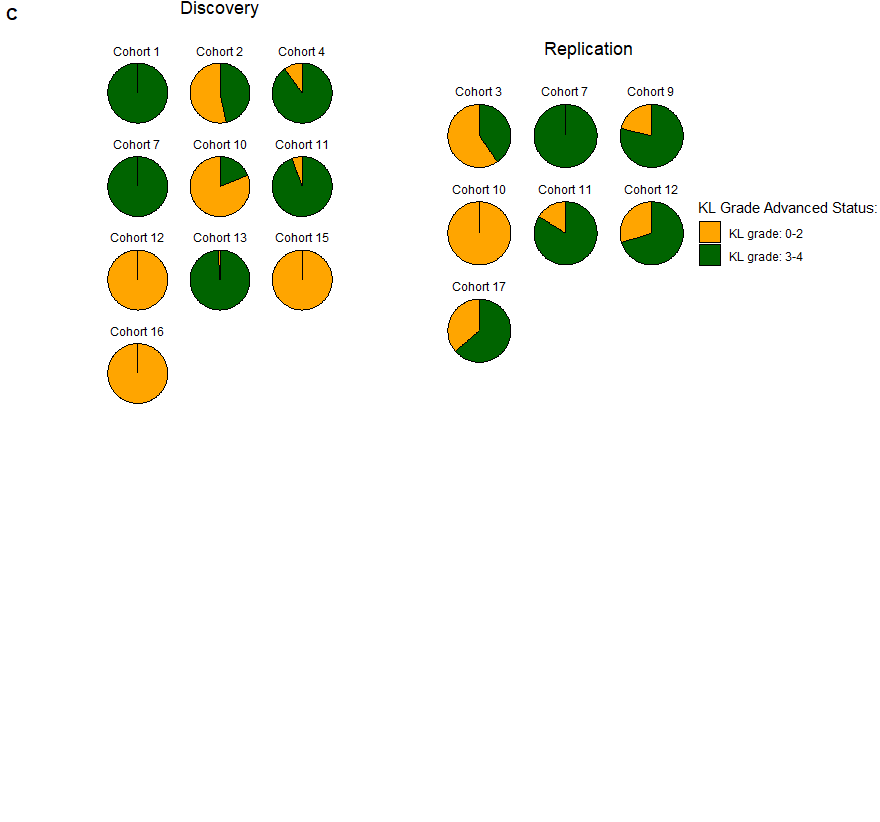

**A**

**C**

**B**

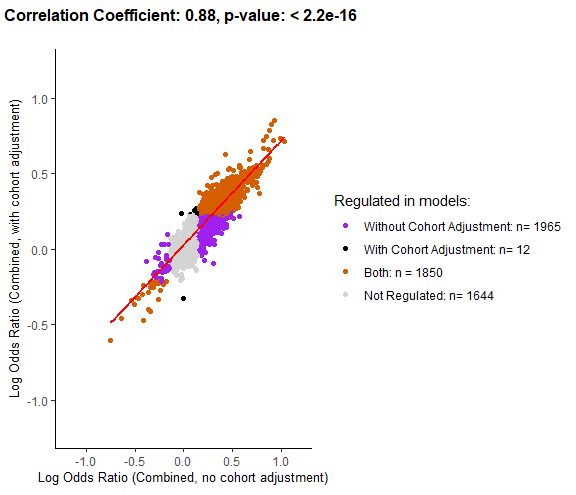

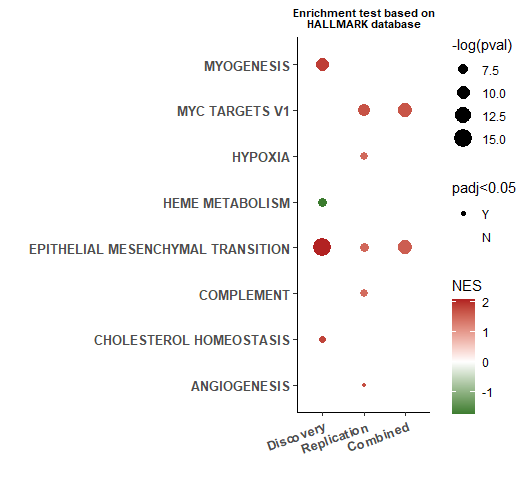

**Supplementary Figure 1**: ***Association between protein abundance and advanced radiographic knee OA status in the non-IPS regressed data* with additional adjustment for cohort.**

Protein abundance was measured in 1,322 samples (Discovery and Replication, spun (N=1,096) and unspun (N = 226)), corrected for spin-status and then adjusted for age, biological sex and cohort (random intercept). **(A)** A scatter plot of log odds ratio from logistic regression models of the association between protein abundance and advanced radiographic disease status using either Combined dataset without adjustment for cohort or with adjustment for cohort (random intercept) is shown with significantly associated proteins in different models shown in different colours (see key). Pearson correlation coefficient and p-value (unadjusted) are presented for the correlation between log odds ratio generated in Combined (non-cohort adjusted and cohort adjusted) analyses. **(B)** Pie charts demonstrating the variation in cohort composition of radiographic disease severity across individual cohorts within Discovery and Replication. **(C)** Bubble plot of significantly enriched pathways (adjusted p-value <0.05) using the Hallmark Gene set for advanced radiographic knee OA status using Discovery, Replication and Combined non-IPS regressed datasets with additional adjustment for cohort (random intercept). Abbreviations: normalised enrichment score (NES), intracellular protein score (IPS). Full list of proteins is available in Supplementary data file 1.

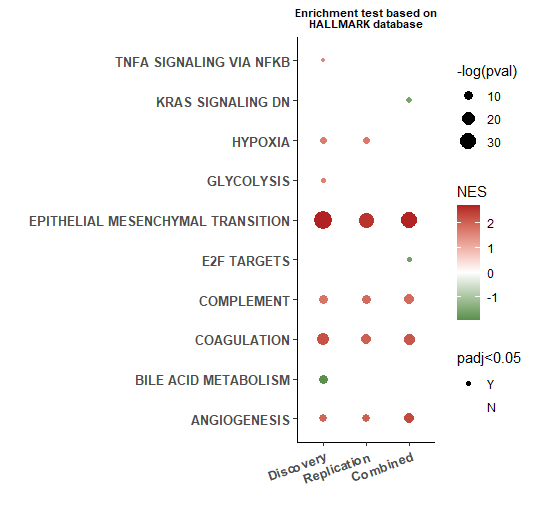

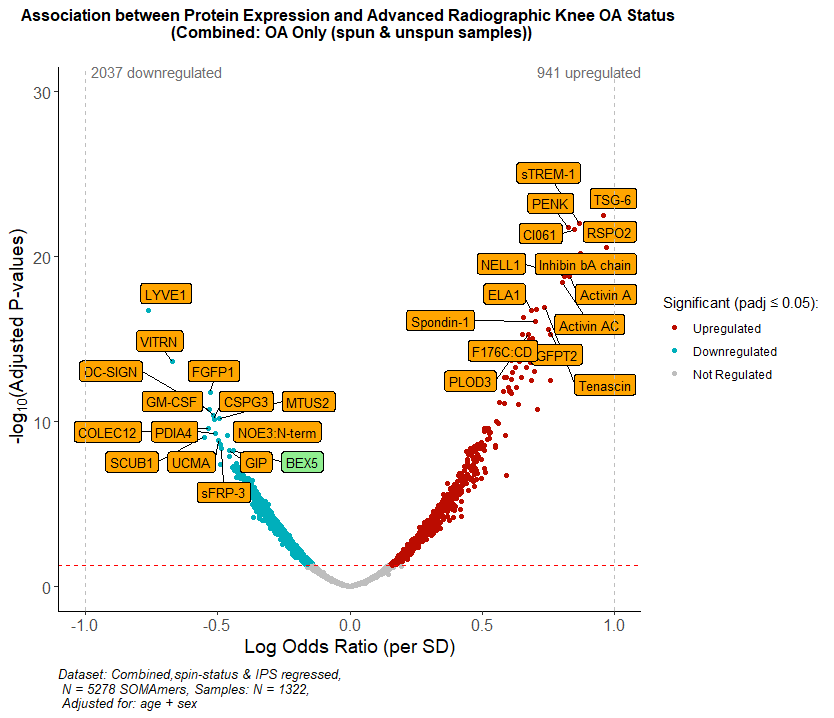

**A**

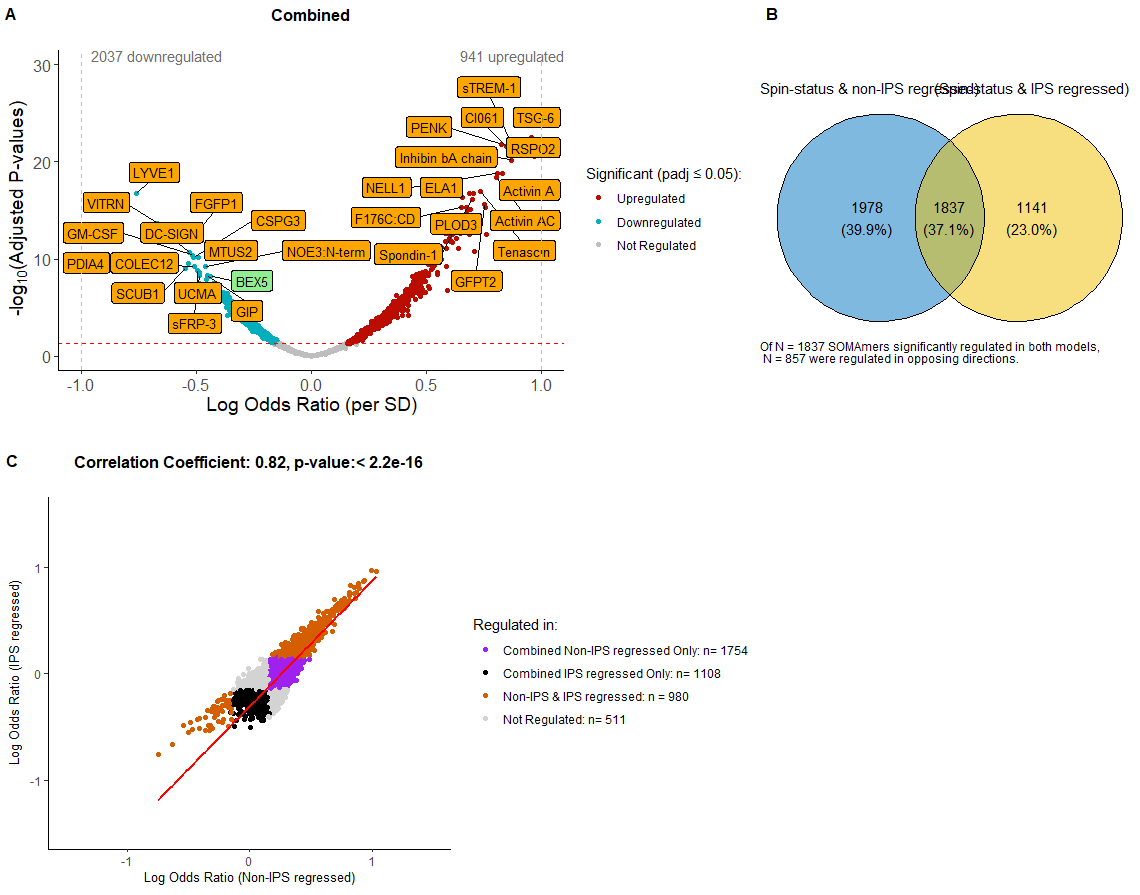

**B**

**C**

**Supplementary Figure 2**: ***Association between protein abundance and advanced radiographic knee OA status in the IPS regressed data.***

Protein abundance was measured in 1,322 samples (Discovery and Replication, spun (N=1096) and unspun (N = 226)), corrected for spin-status and then adjusted for age and biological sex. Additionally, these samples underwent IPS regression. **(A)** Volcano plot showing log odds ratios against adjusted p-values (Benjamini-Hochberg corrected, padj) for proteins associated with advanced radiographic knee OA in the Combined dataset. Proteins in red are positively associated, those in blue negatively associated, with advanced RKOA status (KL grades: 3-4) at an adjusted p-value ≤ 0.05. Top 30 negatively and positively associated proteins by adjusted p-value are labelled. In orange are proteins that replicated (significant at padj ≤0.05 and with effects in the same direction, in Discovery & Replication datasets), and remained significant after Combined dataset was adjusted for cohort (as a random intercept). Proteins that either did not replicate but remained significant after adjustment for cohort, or did replicate but were not significant after cohort adjustment are shown in green. **(B)** Scatter plot of log odds ratio from logistic regression models of the association between protein abundance and advanced radiographic status in non-IPS regressed and IPS regressed analyses is shown with significantly associated proteins in different groups shown in different colours (see key). Pearson correlation coefficient and p-value (unadjusted) are presented for the correlation between log odds ratios generated in non-IPS and IPS regressed analyses using Combined datasets. **(C)** Bubble plot of significantly enriched pathways (adjusted p-value <0.05) using the Hallmark Gene set for advanced RKOA status using Discovery, Replication and Combined IPS regressed datasets. Abbreviations: normalised enrichment score (NES), intracellular protein score (IPS). The full list of proteins is available in Supplementary data file 2.

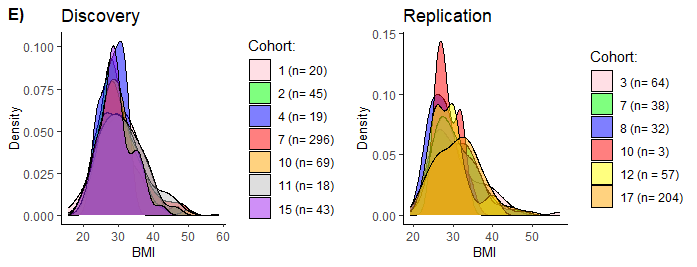

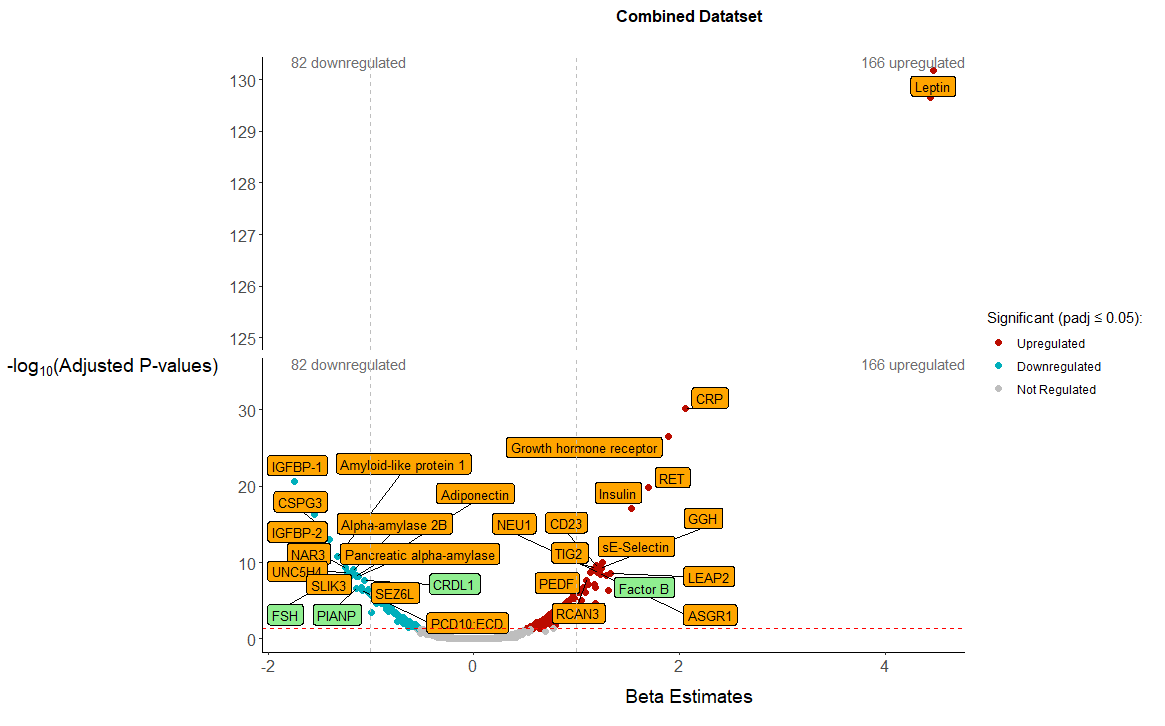

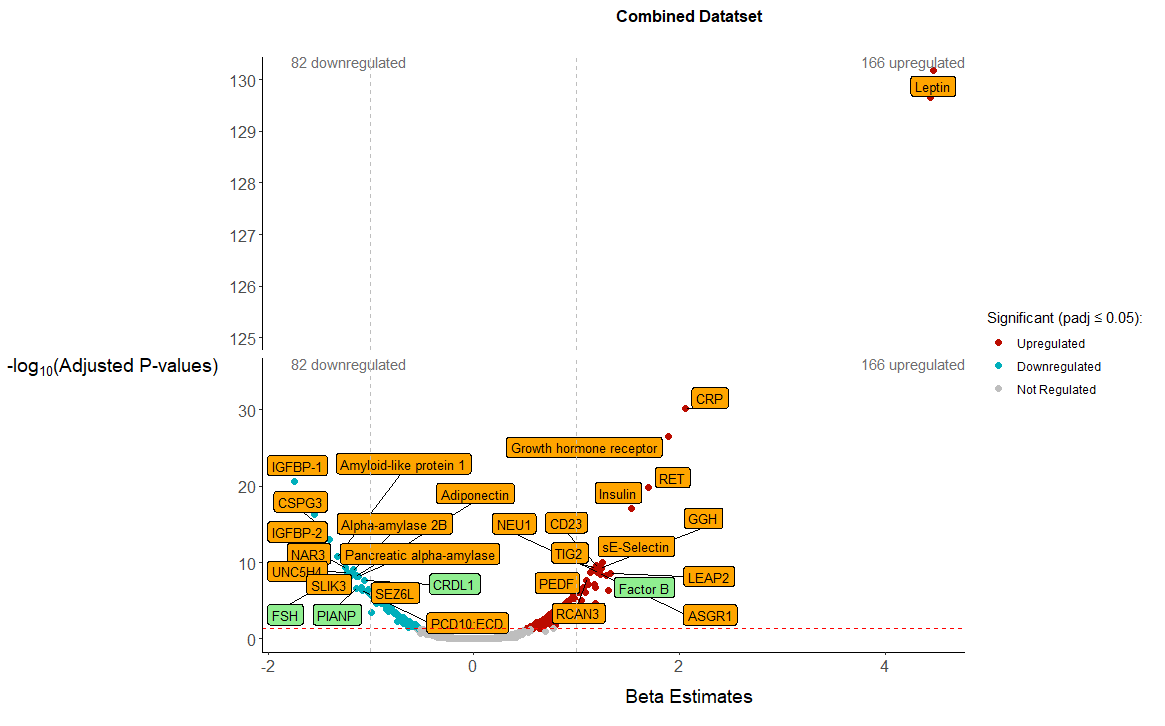

**A**

**B**

**C**

**E**

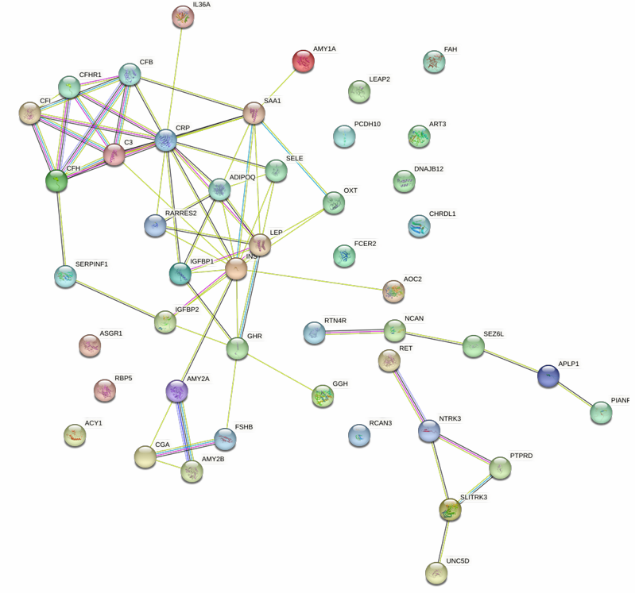

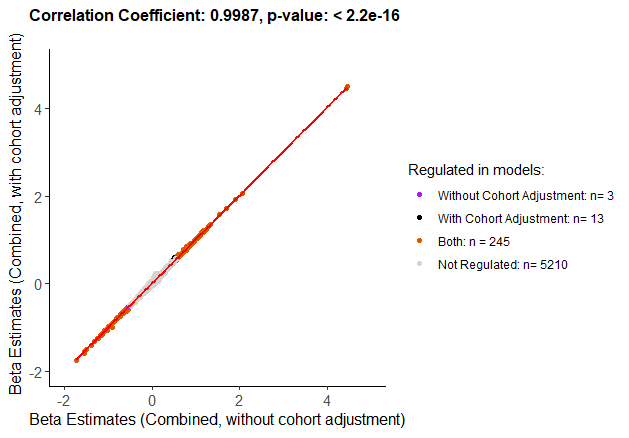

**D**

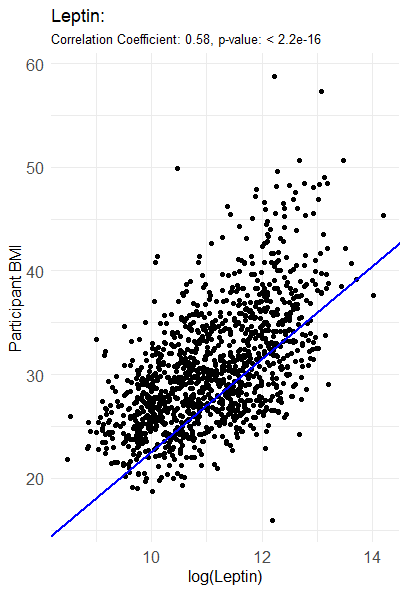

**Supplementary Figure 3**: ***Association between protein abundance and body mass index (BMI) in OA using the non-IPS regressed dataset.***

Protein abundance was measured in 1,236 samples where BMI was available (Combined dataset, spun, N=1,045; unspun, N=191), corrected for spin-status and then adjusted for biological sex and radiographic disease severity. **(A)** Volcano plot showing beta estimates against adjusted p-values (Benjamini-Hochberg corrected) for proteins associated with BMI. Proteins in red are positively associated, those in blue negatively associated, with advanced BMI at an adjusted p-value ≤ 0.05. Top 30 positively and negatively associated proteins by adjusted p-value are labelled. In orange are proteins that replicated (significant at padj ≤0.05 and with effects in the same direction in Discovery & Replication datasets), and remained significant after Combined dataset was adjusted for cohort (random intercept). Proteins that either did not replicate but remained significant after adjustment for cohort, or did replicate but were not significant after cohort adjustment are shown in green. **(B)** Scatter plot of participant BMI against log-transformed leptin protein expression. Pearson correlation coefficient and p-value (unadjusted) are presented, with linear regression line plotted. **(C)** Protein-protein interaction network, using STRING, for those proteins associated with BMI (top 50 most significant based on padj). **(D)** Scatter plot of beta estimates from linear regression models of the association between protein abundance and BMI using the Combined dataset without adjustment for cohort or Combined dataset with adjustment for cohort (random intercept) is shown with significantly associated proteins in different models shown in different colours (see key). Pearson correlation coefficient and p-value (unadjusted) are presented for the correlation between beta estimates generated in Combined analyses with and without adjustment for cohort. **(E)** Distribution of BMI across individual cohorts contained within Discovery and Replication datasets. Abbreviations: body mass index (BMI), intracellular protein score (IPS). Full list of proteins is available in Supplementary data file 3.

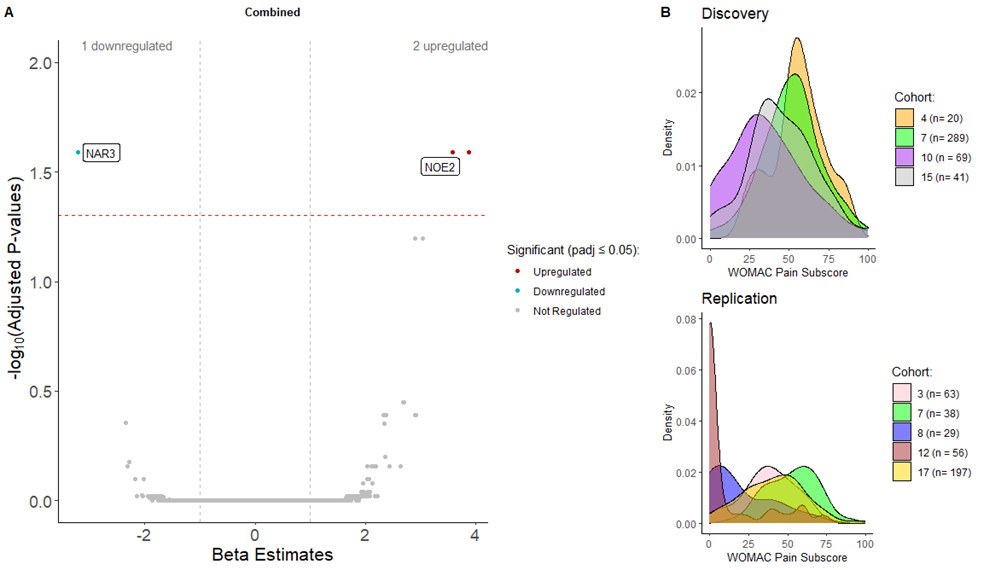

**A**

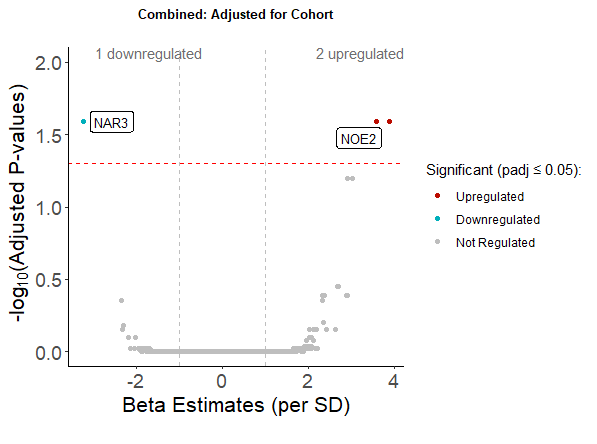

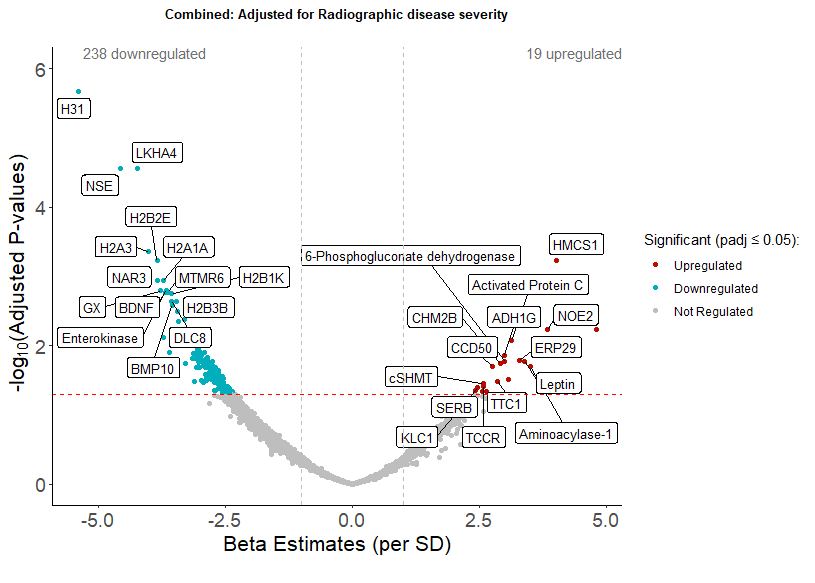

**C**

**D**

**B**

**A**

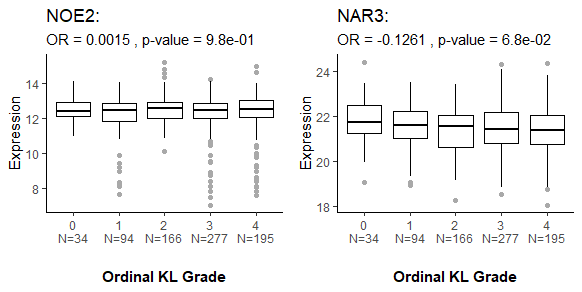

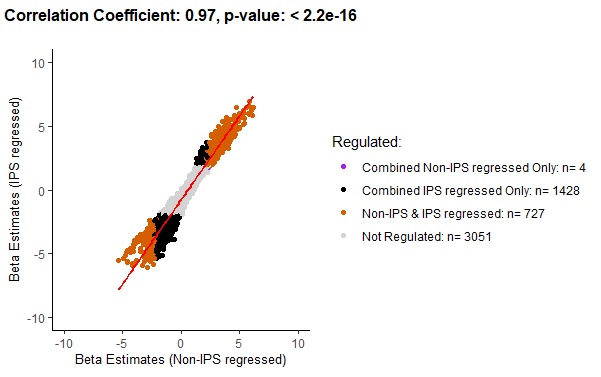

**E**

**Supplementary Figure 4:** ***Association between protein abundance and WOMAC pain subscores in the non-IPS regressed data* *after adjustment for cohort or advanced radiographic disease status*.**

Protein abundance was measured in 805 samples where WOMAC pain was available in the Combined dataset (Discovery and Replication, spun (N=748) and unspun (N = 57)). **(A)** Volcano plot showing beta estimates against adjusted p-values (Benjamini-Hochberg corrected) for proteins associated with an increase or decrease in WOMAC pain subscore in the Combined dataset adjusted for age, biological sex and cohort (random intercept). Two proteins, labelled in white, remained significantly associated after adjusting for cohort (NOE2 was positively associated and NAR3 negatively associated), at an adjusted p-value ≤ 0.05. **(B)** Variable distribution of WOMAC pain subscores is shown for individual cohorts across Discovery and Replication. **(C)** Volcano plot showing beta estimates against adjusted p-values (Benjamini-Hochberg corrected) for proteins associated with increasing WOMAC pain subscore in the Combined dataset, adjusted for age, sex and advanced radiographic disease status. **(D)** Boxplot of NOE2 or NAR3 protein abundance (transformed by natural logarithms) in OA participants using Combined, spin-status corrected, non-IPS regressed data by ordinal KL grade. Associations with ordinal KL grade were tested by ordinal regression analysis (log odds ratio (OR) and unadjusted p-values are presented for each protein for models adjusted for age and biological sex). **(E)** Scatter plot of beta estimates from linear regression models of the association between protein abundance and WOMAC knee pain using the Combined dataset without and without adjustment for IPS is shown with significantly associated proteins in different datasets shown in different colours (see key). Pearson correlation coefficient and p-value (unadjusted) are presented. The full list of proteins is available in Supplementary data files 6 & 7.
